# Quantifying the direct and indirect protection provided by insecticide treated bed nets against malaria

**DOI:** 10.1101/2022.01.21.22269650

**Authors:** H. Juliette T. Unwin, Ellie Sherrard Smith, Thomas S. Churcher, Azra C. Ghani

## Abstract

Long lasting insecticide treated mosquito nets (LLINs) provide both direct and indirect protection against bites from mosquitoes potentially transmitting malaria. Direct personal protection is provided to net users given both the net’s physical barrier and its insecticidal action. Indirect mass protection for the community is afforded through reduced infectious bites per person annually (entomological inoculation rate, EIR). Quantifying these protective effects can help strategize options for net interventions, particularly as insecticide-resistant mosquitoes spread.

These types of protection are inherently linked, rendering it impossible to empirically quantify the contribution of each to the overall ‘community effect’, instead we investigate this with a modelling framework and compare model predictions to trends with Demographic Health Survey (DHS) data.

Our modelling exercise predicts that in a situation with an EIR of 100, the reduction in EIR from an untreated net used by 80% of the population is 52% [95% CI: 12% - 84%] for users and 21% [95% CI: 0% - 57%] for non-users. Due to the impact of the insecticide, the reduction in EIR for LLINs is 89% [95% CI: 67% - 98%] for users and 74% [95% CI: 48% - 92%] for non-users, but this protection reduces as insecticide resistance in mosquitoes increases. Modelled trends in the difference in protection between users and non-users across endemicity and net usage levels are consistent with DHS data (2000-2018).

This study supports the concept of a community effect from LLINs, highlights the value of blocking and killing mosquitoes for community protection. Achieving high LLIN usage is always preferential, but there remains protection to non-users as the number of people using nets increases.

## Introduction

Long-lasting insecticide-treated mosquito nets (LLINs) have been attributed with averting 663 (542– 753 credible interval) million clinical cases (68% of malaria cases) globally across 2000 to 2015 [1]. In recognition of this impact, most malaria-endemic countries in sub-Saharan Africa distribute LLINs universally to communities through mass campaigns operating approximately every 3-years [2, 3]. Adherence to net usage is variable among communities but generally good when access is good [4]. Variable access and usage can be partially explained by uncomfortably high humidity and temperatures in some areas making sleeping beneath nets challenging [5], an absence of perceived risk, mis-information on LLIN utilization [6, 7], and coverage gaps in distribution campaigns [5, 7, 8]. Recently, school-based and health centre ‘top-up’ campaigns are trying to address such coverage gaps [9]. LLINs contain insecticides which kill mosquito vectors so that where net use is not universal, some community protection is never-the-less potentially provided to everyone. The challenge to deliver LLINs universally and the emergence of mosquitoes able to survive exposure to pyrethroid insecticide – the principle active ingredient for LLINs – has led malaria researchers to question the protection offered by insecticide [10, 11]. It is therefore important to quantify the direct and indirect protection offered from LLINs to both users and non-users. We define the indirect protection offered to both users and non-users as the mass community effect. Both the barrier and the insecticide offer direct and indirect protection.

Evidence of the community effect is ambiguous, partly because empirical studies have not been powered to address this question – it would be unethical now to deliberately withhold nets from a cohort of a community. Inferences can be drawn from early studies that empirically tested malaria burden in users and non-users and these tend to show intuitively that burden is lower in the cohorts using nets and some parallel but smaller reduction is often seen in non-user cohorts [12–15]. Evidence against a community effect is sometimes cited from a study done in The Gambia [16] because malaria prevalence was higher among non-users living within village clusters of people using nets than within villages without nets. However, the baseline prevalence varied considerably across the village clusters (ranging 28.7 – 71.2% prevalence in children 1 to 4 years of age) and this alone may explain the observed results. Modelling studies have also addressed this question [17–20] but inherent within models are the assumptions that they make, so modelling evidence is generally considered to be poor, particularly when empirical data are not available to corroborate conclusions.

The community effect is also an ambiguous metric to define. A mosquito net will provide direct protection to the individual who uses a net based on the physical barrier directly preventing bites. It can provide added protection to that individual through the killing action of insecticide, as fewer mosquitoes are then present to bite during the hours of the day when the net is not in use. As the proportion of people in a community using nets increases, this protection remains but is added to by: i) the physical barrier that still prevents bites directly to the user; ii) the killing action that reduces the number of mosquitoes across the community so too reducing any bites to the users and non-users of nets, and iii) the consequential reduced transmission because fewer infectious bites will occur for those still receiving bites. Clearly, the interconnectivity of these types of protection renders it impossible to empirically quantify the contribution of each to the overall ‘community effect’, but this can be addressed with a validated modelling framework.

Here we compared the difference in prevalence between users and non-users of LLINs in Demographic Health Survey (DHS) data and predicted estimates from an established mechanistic transmission model for *Plasmodium falciparum* [21–24] for multiple administration units across Africa. We then use the transmission model to tease apart the direct benefit of LLINs from the mass community effect, which is not possible to do from trial data, and investigate what happens as mosquitoes show increasing resistance to pyrethroid insecticide.

## Methods

### Demographic health survey data comparison

We use Demographic and Health survey (DHS) data to evaluate the difference in prevalence in 6-59-month-olds for clusters with different prevalence and usage levels. We include all available DHS data for African countries since 2010 with geolocated cluster data, *Plasmodium falciparum* malaria prevalence and LLIN usage [25]. In these DHS data, a cluster represents a collection of neighbouring villages using a small variable number of health facilities for clinical testing and treatment, the total number of people of all ages in each cluster varies from 40 to 543. Net usage is estimated from the survey by asking the heads of households if individuals of all ages slept beneath a LLIN the previous night. A total of 47 surveys from 21 countries are included; they are listed in Table S2. We use the *rdhs* package to extract data [26]. We only consider usage and RDT outcomes from clusters where 5 or more children use a net and 5 or more children do not use a net, which results in the inclusion of 115,234 individuals 6-59-months-old in 5340 clusters.

We aggregated our data using survey weights to the first administrative level (often the District or Province level of a country) to investigate if these trends were significantly driven by resistance using a linear regression. We do not have data from susceptibility bioassays (or other metrics for approximations of pyrethroid resistance) matched by date and location for the DHS data and found, in preliminary analyses, no temporal association between the difference in prevalence of users and non-users holding country as a random effect (Linear mixed effects model fit with REML, time: *p* = 0.186, R package: lme4 [27]) illustrating the challenge of using these data to explore the impact of resistance. Instead, we use a systematic review of susceptibility bioassays collated from 2000 – 2018 across the African continent [28, 29]. Assuming an increasing logistic function over time for percentage-survival at bioassay testing, these data provide an estimate of the level of resistance for each region at the District or Province scale (Lambert et al in prep). Clearly, there are limitations with this broad-brush approach, not least given the variability in both the assay and geographic expressions of resistance [30–32] – we present these limitations in the discussion – but we include the analysis to see whether there is a signal in the empirical data, and to give some context for our subsequent modelling exploration of what increasing resistance may mean for continued protection offered by a community effect.

### Modelling approach

The difference in user and non-user prevalence from the DHS data are compared to model predictions from a deterministic version of a well-established compartmental *Plasmodium falciparum* malaria transmission model [21]. This model incorporates a full dynamic mosquito-vector component [22], which satisfactorily represents vector-control interventions [33]. We briefly describe the model below focusing on how the effect of LLINs are incorporated, but the full mathematical detail is provided in Supplementary Information 1 and the following references: [21, 23, 24].

The community considered is split into age, heterogeneity and intervention compartments. In this model we consider two intervention groups, those who sleep under an LLIN (LLIN users) and those who do not (non-LLIN users). The population is initially modelled as susceptible, with people moving to the infected state at a given rate through bites from infectious mosquitoes. Following a period reflecting liver-stage infection, a proportion of the individuals in the infected compartment become symptomatic and seek treatment. The proportion who receives successful treatment experience a period of drug-dependent prophylaxis before returning to the susceptible compartment. The proportion of symptomatic individuals who do not receive treatment experience a period of symptomatic disease before transitioning into the asymptomatic compartment. The proportion of the population who are asymptomatic move from patent infection to sub-patent before natural recovery transitions them back to the susceptible compartment. We also include superinfection in the model from asymptomatic and sub-patent states. Superinfection is defined here as an infected person who can be bitten and infected again by an infectious mosquito.

We consider three types of immunity in our model, with parameters estimated by fitting to clinical incidence and parasite prevalence data [21, 34]. Clinical immunity is exposure-driven, so age dependent, and protects the population against clinical disease. Antiparasitic immunity develops through age and exposure to infection and reduces how easily infections are detected through the control of parasite density. Anti-infection immunity develops later in life and reduces the probability that an infectious bite results in patent infection. All human infection states are assumed to be infectious to mosquitoes, with infectivity correlated with parasite density.

The compartmental vector model has larval stages and adult female mosquitoes similar to Griffin *et al*. [21] and White *et al*. [22]. This results in the following entomological inoculation rate (EIR),

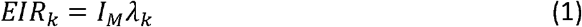

where I_M_ is the number of infectious bites and *λ*_*k*_ is the intervention compartment-dependent biting rate, which varies depending on the LLIN usage and pyrethroid resistance values (k=1 for non-users and 2 for users). The probability of a blood-seeking mosquito successfully feeding depends on the behaviour of the mosquito and the anti-vectoral defences employed by the human host population. There are three possible outcomes tracked in the model once a mosquito enters a house to feed: it can repeat (*r*_*N*_), feed successfully (*S*_*N*_) or die (*d*_*N*_). Over time, the efficacy of the repellence effects of the LLIN fluctuates from a maximum, *r*_*N*0_, to a non-zero level, *r*_*NM*_, which reflects the protection still provided by a net that no longer has any insecticidal effect (and potentially some holes). The killing effect of ITNs decrease from a maximum *d*_*N* 0_ when the insecticide is working optimally. The outcomes *r*_*N*_, *d*_*N*_ and *s*_*N*_ always sum to 1.

The net model has been parameterised using data from a systematic review of experimental hut studies [35]. This review collated 90 experimental hut trial arms with an untreated net included as the control. These data are used to parameterise the benefit of an untreated net relative to a no-net control adopting the same method as LLINs. This is important because untreated nets are also thought to increase mosquito mortality relative to a no-net control, so prescribing all mortality due to the action of the insecticide is likely to over-estimate insecticidal impact. Briefly, the entomological impact of LLINs is tested in huts to estimate the probability that a mosquito will successfully blood feed, be killed, or continue on without feeding (a combination of deterrence; not entering the LLIN hut at all, and repellence; entering and then exiting without feeding). In the absence of a mosquito net, it is assumed that 69.9% of mosquitoes will successfully feed [36, 37] and otherwise exit. The efficacy of the killing effect measured from LLIN experimental hut trials is associated with mosquito mortality in the discriminatory dose susceptibility bioassay tests that are used to approximate any changes in pyrethroid resistance over time for a given location. The associations between experimental hut mortality, successful blood-feeding and deterrence, that are driven by LLIN presence, are also predictably altered as the proportion of mosquitoes surviving a susceptibility bioassay test increases [35]. Therefore, these associations can be used to estimate the lost impact of LLINs in the presence of pyrethroid resistant mosquito populations. Net use is assumed to be random within the population and we assume systematic use of nets over time (LLIN-user sleep under LLINs every night).

When comparing the model data to the DHS data, we used bisection to fit the initial EIR (model input number) to match the post intervention prevalence of 0.1 to 0.8 in children aged 6-59-months-old for LLIN usage levels of 10% to 80%.

### Direct vs indirect (mass community) protection

Since it is hard to address the different types of protections offered to users and non-users from data, we used our transmission model to tease apart the direct and indirect (mass community effect) protection offered by LLINs. In addition to a control where nobody in the community is given any nets, we identify 4 scenarios that describe the different types of protection offered by untreated nets or LLINs to users and non-users (Figure 1). We fix the EIR over time in some scenarios (by breaking the link between current malaria endemicity and the human force of infection) to disentangle the direct and mass community effect of nets. This is implemented in the model by fixing the number of infectious mosquitoes (Equation (1)) to the number in the control and fixing the biting rate of non-users (k=1) to the biting rate in the control scenario.

**Figure 1:**
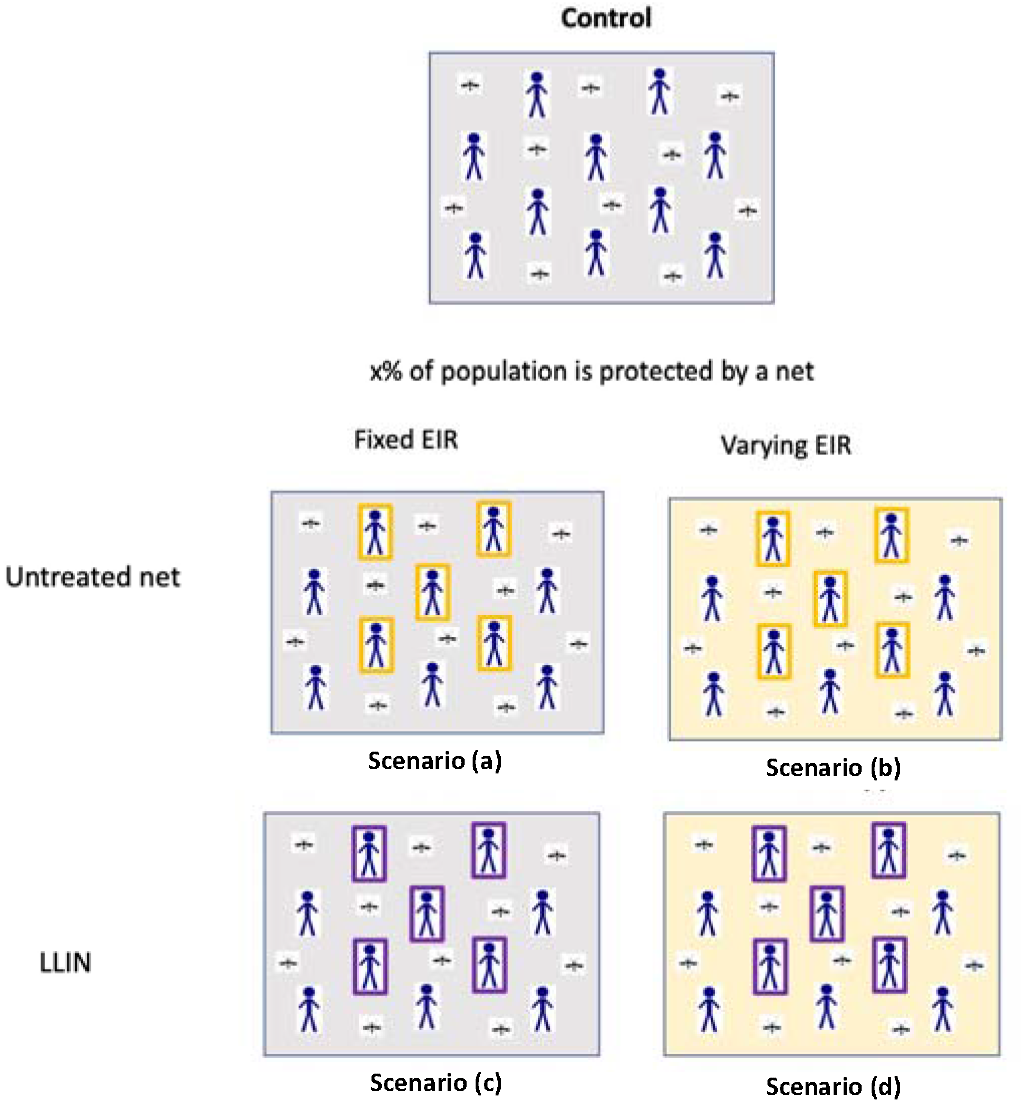
A schematic of control and 5 different theoretical scenarios. Top panel: Nobody uses a net in the control scenario, and this is used as the counterfactual to compare the other scenarios against. Bottom panels: Scenario (a) illustrates direct protection from the barrier to a proportion of the population, in the analysis we consider an individual (1/100,000),10%, 50% or 80% net use. If x is 50% as illustrated, 50% of people consistently used an untreated net (orange rectangle) for a fixed EIR (i.e. the entomological inoculation rate, EIR, remains constant irrespective of the net use, grey box). Scenario (b) illustrates direct + indirect protection from the barrier to a percentage of the population. This time we illustrate that 50% of people are protected with an untreated net (orange rectangle) for a varying EIR (i.e. EIR varies over time according to the net use as estimated by the mathematical model, yellow box). Similarly, scenario (c) illustrates direct protection from the barrier and insecticide to a percentage of the population via an insecticide treated net (purple rectangle). Scenario (d) illustrates direct + indirect protection from the barrier and insecticide to a percentage of the population.

a. *Direct protection from barrier to x% of the population. x% of* people protected with an untreated net for a fixed EIR.
b. *Direct + indirect protection from barrier to x% of the population. x% of* people protected with an untreated net for a varying EIR.
c. *Direct protection from barrier and insecticide to x% of the population. x% of* people protected with a LLIN for a fixed EIR.
d. *Direct + indirect protection from barrier and insecticide to x% of the population. x% of* people protected with a LLIN for a varying EIR.

We illustrate the impact of the mass community effect by investigating the reduction in both EIR and all-age prevalence from the control scenario for users and non-users. We repeat scenarios (a) - (d) for four different usages: an individual (1/100,000), 10%, 50% and 80% (the maximum usage expected in a community – that is, the proportion of people using a mosquito net at deployment immediately after the mass net distribution) to investigate how the mass community effect varies with usage. In each scenario, we run the model for one year and then add the intervention. Initially, we subtract the average of the EIR/prevalence between years 3 and 6 from the control (to average over the three-year waning cycle) so we are presenting a reduction in EIR/prevalence due to the intervention. We then calculate the reduction in the indirect part of the protection offered by subtracting the direct protection from the direct + indirect protection, for example the indirect protection from the barrier for an individual is scenario *control – (b) – (control – (a))* or *(a) – (b)*. Further, we calculate the additional benefit of the insecticide by subtracting the protection from a scenario without insecticide from the similar scenario with insecticide, for example the additional direct protection from insecticide to an individual is *control – (c) – (control – (a))* or *(a) – (c)*, see Table 1.

**Table 1:** Equations for calculating reduction in EIR and prevalence from model runs. These refer to the scenarios presented in Figure 1 and are based on a deterministic model of malaria transmission.

|  | Direct | Indirect |
| --- | --- | --- |
| **Barrier** | control – a | a – b |
| **Additional protection from insecticide** | a – c | b – d – (a – c) |

We present uncertainty in our model estimates by varying the following important parameters: the proportion of mosquitoes that repeat and the proportion of mosquitoes that die for LLINs and untreated nets and the proportion of bites taken on humans in bed (Table S1, Supplementary Text 1). Other parameters that influence both users and non-users, such as the time spent by a mosquito looking for a blood-meal, are kept constant due to the modelling framework. There is a wide range of parameter uncertainty for the efficacy of the different nets and how LLINs are influenced by pyrethroid resistance. Here we assume that the efficacy of LLINs can be no worse than untreated nets. This assumption should be verified with naturally aged nets from the field, as different net brands might physically deteriorate at different rates [38], though in the absence of data on brands of nets used in various locations, this assumption seems the most parsimonious. Uncertainty in our LLIN parameterisations is carried through from the analysis of empirical data studies using experimental huts [32]. One thousand posterior parameter draws are taken from statistical fits to the empirical data measuring mortality, successful feeding, and deterrence [32]. The data are collated following Griffin [21] to provide ranges for uncertainty in net efficacy parameters (Table S1). The median and 90% credible intervals of this range are then included for the sensitivity analysis. We investigated how the direct and mass community effect of LLINs might change with increasing levels of pyrethroid resistance in the local mosquito population.

## Results

### Comparison of model outputs to DHS data

We illustrate the process we take to decouple estimates of prevalence in users and non-users of mosquito nets in Figure 2A for both treated and untreated nets (see Figure S1 for net parameterisation decay through time assuming a three-year net distribution cycle). For a population of 100,000 people, we simulate LLINs being used by 50% of the population at random from year 1 and show the all-age prevalence across the whole community falls from 60% to approximately 40% by year 2 for treated nets and 56% for untreated nets. In the example, it is clear the assumptions of the model indicate that most of this protection is afforded to net users – reducing all-age prevalence of insecticide-treated net users to 33% - whilst there is less reduction in the non-user cohort for the treated net scenario (all-age prevalence of non-users is reduced to about 47%). The reduction in prevalence predicted by the model varies depending on the EIR, but some protection is offered to non-users by having any nets in the community (Figure 2B).

**Figure 2:**
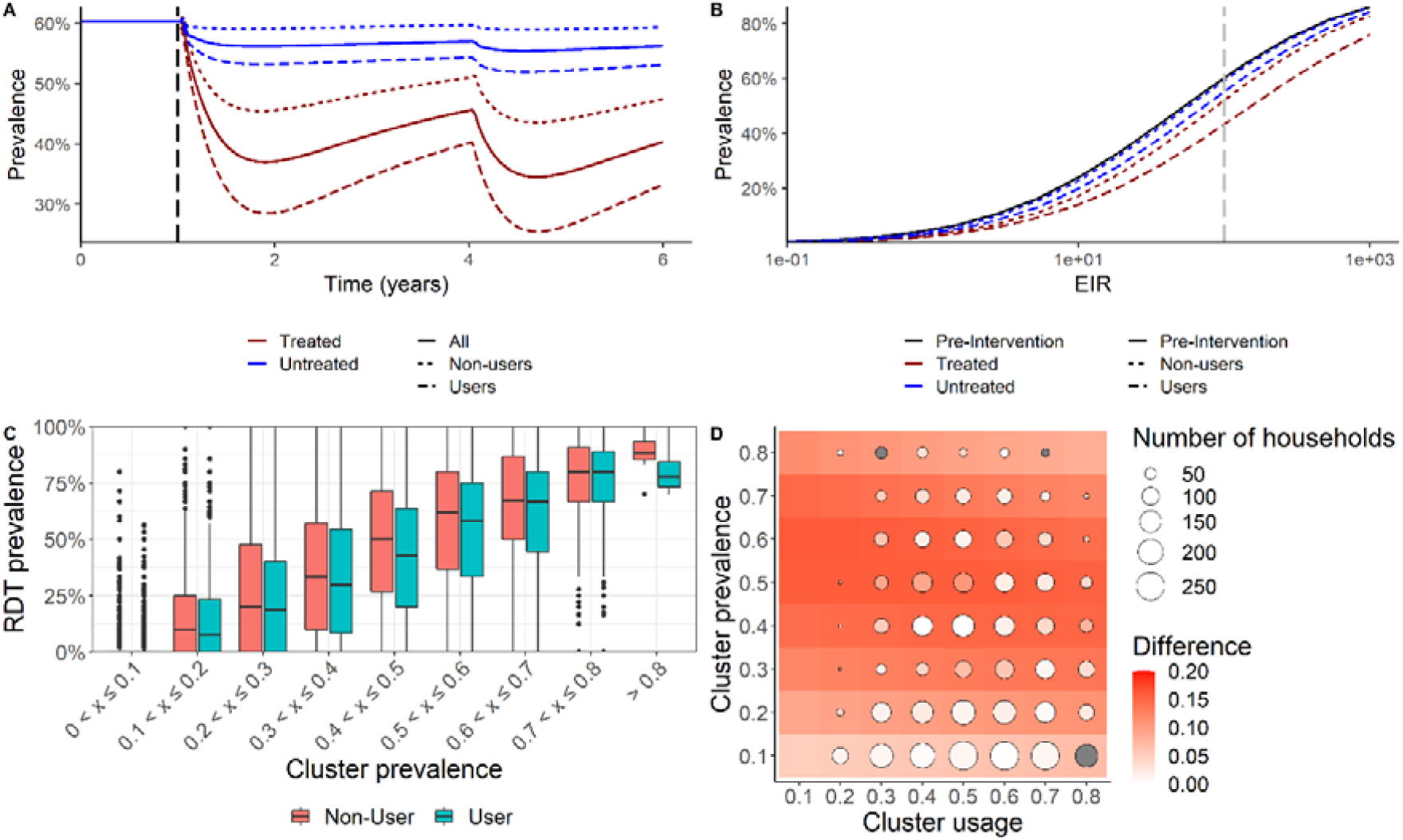
Protected impact on malaria prevalence of standard pyrethroid long-lasting insecticide treated nets (LLINs) for net users and non-users. (A) All-age malaria slide prevalence for a perennial setting with an initial entomological inoculation rate (EIR) of 100 bites per person per year. At year 1 (indicated by the vertical dashed black line), in this example, 50% of the population switch to using LLINs. (B) The dependency of all-age malaria parasite prevalence on the annual EIR for the baseline pre-intervention scenario (black solid line), the LLIN users (blue dashed line), and non-users (red line). The EIR of 100, which is the simulation shown in A, is indicated by the vertical dashed line. (C) RDT prevalence from DHS surveys for users (blue) and non-users (red) for different cluster prevalences. (D) The absolute difference in prevalence between LLIN users and non-users aged 6-59-months from the DHS survey (points, size represents number of data points) and model estimates (tiles). Squares with no point represent cluster usage prevalence combinations that were missing from the data and grey circles show where a negative difference was obtained. We only display data where more than 25 households were included in the cluster.

Figure 2C shows the prevalence between users and non-users from the DHS data for overall prevalence clustered into 10% prevalence bands and we see there is only a small difference between users and non-users in each band. To explore whether our model structure reflects the empirical evidence, the DHS data, categorised by prevalence in children 6 to 59 months of age and the level of net usage among these cohorts, is overlaid on the matrix plot in Figure 2D. The trends match, supporting the hypothesis that any difference in malaria burden in users and non-users is not influenced by usage but more likely associated with the local endemicity of a setting. There is little variation in the difference in the field data between users and non-users for a given prevalence compared to the model suggesting current model structure may be exaggerating the difference. In both models and data, the absolute difference in prevalence peaks for moderate 50% prevalence levels.

We did not detect a signal from the DHS that the difference between users and non-users changed over time or with the level of pyrethroid resistance. This is perhaps unsurprising given that we use averaged data for large geographic regions to assign resistance status to small DHS sample points (areas with 40 – 543 people). The model estimates the impact of resistance on the difference between users and non-users is relatively modest (Figure S2) which supports the lack of a clear trend in the DHS data.

### Modelling exploration of types of protection from nets

In the absence of data, we use the model to intuit the reduction in EIR (Figure 3) that is attributable to the different types of direct and indirect protection provided by mosquito net use. In the first instance, this analysis assumes that no mosquitoes are resistant to the insecticide used on the nets.

**Figure 3:**
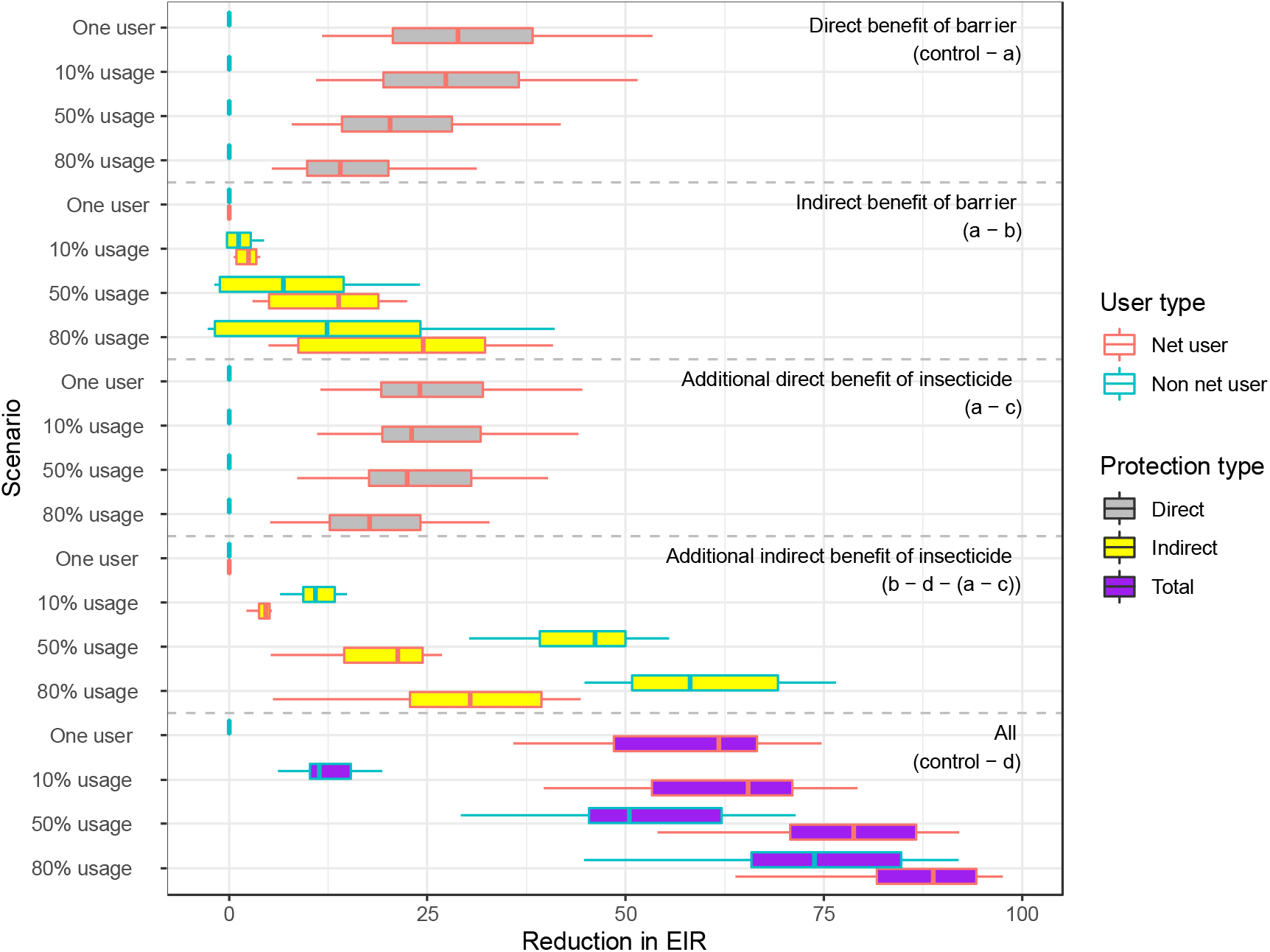
Reduction in EIR from direct and mass community (indirect) protection offered by mosquito nets for a pre-intervention EIR of 100. The reduction in EIR is calculated relative to a control scenario, where nobody in the population is given a net, for the 8 scenarios detailed in the methods section. Scenarios (a) – (d) are repeated for an individual using a net and 10%, 50% and 80% of the population using nets. The reduction for users is shown in red and for non-users is shown in blue. Direct reductions in EIR are filled in grey, indirect reductions in yellow and total reductions in purple.

### Direct benefit of barrier

First, we consider how the direct protection offered by a barrier reduces the EIR at different levels of mosquito net usage. In scenario (a) there is no insecticide on the net, so the protection provided by the barrier is assumed the same as to users of untreated nets (Figure 1). In our definition, the barrier does not influence non-users, regardless of usage, so EIR remains unchanged. When one user in the population is protected, the direct impact from the barrier to that one user is a reduction in EIR of 29% [95% confidence interval (CI): 13% - 53%]. Under the assumptions of the model, any individual user would be protected by this level of reduced EIR.

However, the level of EIR reduction from the barrier to a net-user does vary with usage level. As the usage increases, the direct protection from the barrier to those using an untreated net progressively reduces to a reduction in EIR of 14% [95% CI: 6% – 31%] at 80% usage. This is due to the repellence action of the untreated barrier, which causes the number of mosquito bites on users to increase as more people use nets. When there is only one net user in the population, they will be highly unlikely to receive any bites from mosquitoes which have attempted to feed elsewhere. As usage increases more mosquitoes will attempt to bite on users that have previously tried to bite on other users. As protection from untreated nets is partial, the EIR on users will therefore increase, reducing the direct benefit.

### Indirect benefit of barrier

Intuitively, the model predicts that indirect benefits from only one net in the community (scenario (a) – (b)) are minimal but that the magnitude of this type of protection increases with usage to a reduction in EIR of 24% [95% CI: 5% - 41%] at 80% usage for users and 12% [95% CI: -3% - 40%] for non-users. As usage increases, in general, more mosquitoes are killed, so the median reduction in EIR for each non-user increases. However, there is a chance that the indirect benefit to non-users may be negative under scenarios where very few mosquitoes are killed. Greater indirect benefits are seen in users rather than non-users because, as net usage increases, net-users do not experience an increase in mosquito bites caused by mosquitoes being dissuaded from feeding on users (conversely, non-users will receive more bites as net use increases as more mosquitoes will be repelled from users).

Figure S3 illustrates the combined direct and indirect benefit of the barrier alone (the same as an untreated net). It indicates that the increase in protection from the indirect benefit of the barrier at higher usage levels to users outweighs the decrease in direct protection from the barrier at higher usages. Therefore, the total protection (direct + indirect) from a barrier in users increases with usage: it is a reduction in EIR of 29% [95% CI: 12% - 53%] for one individual using a net which increases to 42% [95% CI: 11% - 71%] when 80% of the population use a net. This corresponds to no meaningful reduction in EIR when just one individual uses a net to 12% [95% CI: -3% to 40%] for non-users (when 80% of the population are using nets).

### Direct benefit from insecticide

The addition of insecticide on LLINs increases the mosquito mortality and reduces the probability of repeating a feeding attempt. We isolate the additional benefit of this insecticide from the direct benefit of the barrier by subtracting the impact from scenario (c) from the respective scenario (a) (Figure 1 & 3). The direct benefit of insecticide does not impact non-users (blue bars) but reduces for users (red bars) with increasing usage (the reduction in EIR for that individual is 24% [95% CI: 12% - 44%] when only one individual has a LLIN to 18% [95% CI: 6% - 32%] when 80% of the population use LLINs). This is again caused by the deterrent/repellent nature of the insecticide which dissuades mosquitoes from entering houses and forces them to feed elsewhere. As usage goes up the number of mosquitoes attempting to enter houses increases (as less bites are successful) though the relationship with usage is less pronounced due to the killing actions of LLINs.

### Indirect benefit from insecticide

Finally, we see that the indirect additional benefit of insecticide increases with usage for both user and non-users. The indirect impact of insecticide mostly increases the reduction in EIR, as expected, due to the killing action of the insecticide. This is minimal when only one user in the population has a net (reduction in EIR for both users and non-users is 0% [95% CI: 0% - 1%]), but further reduces the EIR by 30% [95% CI: 7% - 44%] in users and 58% [95% CI: 45% - 76%] in non-users when 80% of the population sleeps under a net. The larger indirect protection from the insecticide to non-users rather than to users is because users are already protected through the direct benefit of the barrier and the insecticide.

### Combined benefit from LLINs

The overall protection provided to a single person using an LLIN is estimated reduction in EIR of 62% [95% CI: 36% - 74%] (sum of the above four different types of protection). This is ∼77% larger than the model predicts for a single person using an untreated net, which is predicted to reduce their personal EIR by 35% [95% CI: 12% - 64%] (Direct + indirect protection from barrier Figure S3). Protection provided to users and non-users of LLIN and untreated nets increases as their use increases in the community. When usage increases to 80%, the reduction in EIR for LLINs is 89% [95% CI: 67% - 98%] for users and 74% [95% CI: 48% - 92%] for non-users. In contrast, the total protection offered by an untreated net used by 80% of the population is 52% [95% CI: 12% - 84%] for users and 21% [95% CI: 0% - 57%] for non-users (Figure S3). It is interesting to note that the model predicts ∼20 % more protection is provided to people not sleeping under a net in a community with 80% LLIN use than would be provided by a single LLIN user in a community of non-users.

The specific results presented in Figure 3 are for an initial pre-intervention EIR of 100. Absolute estimates and the relative difference between net types and users/non-users will vary with endemicity (baseline EIR) though similar trends are observed (Figure 2B, Figure S4). The relative magnitude of the differences will also vary in low transmission settings as some intervention may cause local elimination. We see that the relative reduction in EIR is on average slightly higher for an initial EIR of 10 than an initial EIR of 100. In addition to EIR, we are also able to tease apart the protection offered to users and non-users through the resulting reduction in all-age prevalence (Figure S5).

### Types of protection from LLINs in the context of pyrethroid resistance

Lastly, we consider how these four components (direct benefit of barrier, indirect benefit of barrier, direct benefit from insecticide and indirect benefit from insecticide) vary with increasing pyrethroid resistance in the local mosquito population (Figure 4). The direct and indirect benefits of the barrier do not change with resistance because the barrier mode of action is not impacted by pyrethroid resistance. We see that as pyrethroid resistance increases the additional direct impact of the insecticide for LLIN-users decreases as does the additional indirect benefit of the insecticide for LLIN-users and non-users. For example, at no resistance and 50% usage, the additional direct benefit of the insecticide for a LLIN-user is a reduction in EIR of 22 [95% CI: 9 - 39], but this falls to 15 [95% CI: 6 - 26] reduction in EIR at 80% resistance. In addition, at the same usage, the additional indirect benefit of insecticide is a reduction in EIR of 21 [95% CI: 6 - 27] at 0% resistance for users and 46 [95% CI: 31 - 55] reduction for non-users, but this falls to a reduction in EIR of 15 [95% CI: 9 - 17] and 26 [95% CI: 16 - 35] respectively at 80% resistance. Similar patterns are seen between users and non-users for different levels of LLIN usage. Overall, the relationship between the level of resistance and protection is non-linear, with greater reductions seen at higher levels of resistance in the local mosquito population (Figure 4, Figure S6). Again, the trends are similar for other initial EIRs (Figure S7) and for all-age prevalence (Figure S8).

**Figure 4:**
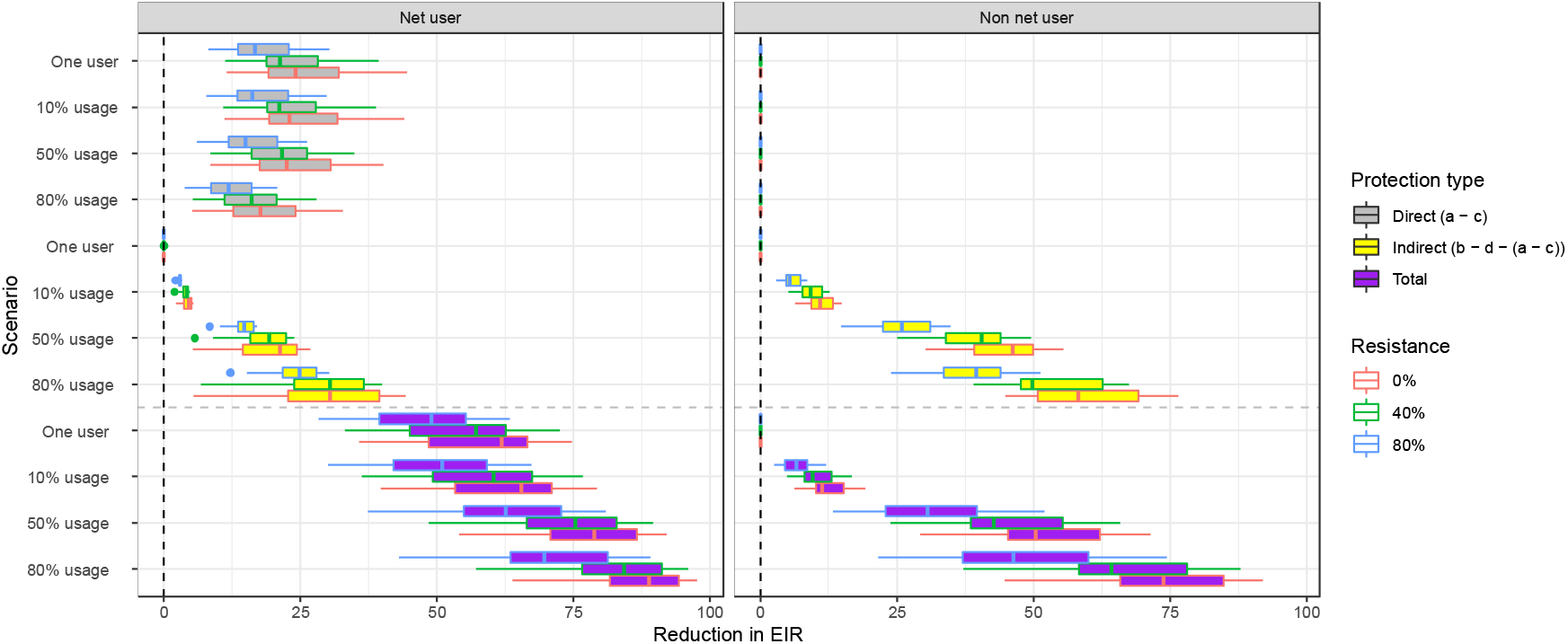
Reduction in EIR from mass community protection offered by LLIN at differing levels of pyrethroid resistance for a pre-intervention EIR of 10. Left figure shows the effect for users and right figure shows the effect for non-users. The reduction in EIR is calculated from a control, where nobody in the population is given a net, for the scenarios detailed in the methods section where insecticide is included. Usages of 1 person in the population, 10%, 50% and 80% are considered for 0% resistance (as in Figure 3) and 40% and 80% resistance. The barrier only effect remains constant at differing levels of resistance so are not included.

## Discussion

Our results suggest that LLINs provide substantial protection to users and non-users through both the physical barrier and the mass community effect and indicate that the role that LLINs play in reducing mosquito populations cannot be underestimated. We use DHS data to explore how well the mechanistic model can reflect observations from the field. These data report rapid diagnostic test (RDT) results and whether individuals slept beneath nets the previous night. Taking these data at the lowest possible spatial scale (clusters), we can see that LLIN users are predicted to have lower prevalence in 6-59-month-olds than non-LLIN users from matched clusters (Figure 1C). This result is consistent with recent studies that saw similar patterns between users and non-users [39–41].

The deterministic model we employ tends to overestimate the difference in prevalence in LLIN users and non-LLIN users, though the overall trends – that the difference is biggest for a cluster prevalence of 50% across all usage levels, tailing away at lower or higher endemicities – are broadly captured. This difference is likely because of the innate variability in real world data resulting in less clear trends; we do not know the local ecologies for each DHS cluster but we do know these can be highly variable e.g. [31, 42, 43]. This can be seen in Figure 2C where the boxplots show the clear variation in prevalence between users and non-users in the different clusters. Some of this variation will be exacerbated by sampling bias as some clusters being relatively small (with 20% of clusters have between 10 and 15 children). The modelling framework used in this analysis is unable to account for this sampling bias which would be better examined using an individual-based model. Our model also assumes systematic use of LLINs over time, i.e. users will always use a LLIN whilst non-users will not. In reality, use will vary from night to night, which reduces the difference in malaria burden between users and non-users. There is currently no method of assessing this variability from the DHS data which simply asks whether a child slept under a net the previous night. In addition, our model also assumes LLINs are distributed at random within the community, whereas people maybe more likely to use nets in regions of the cluster more prone to mosquito biting (for example, nearer breeding sites), further minimising the difference between users and non-users. Seasonality is also ignored given the complexity to untangle the direct and indirect types of protection afforded by nets. On this basis, our model makes assumptions on fixed parameter estimates (Table 2) that restrict cohorts to constant levels of exposure in the presence of mosquito nets throughout the year – although we explore a range of values in repeated model simulations to better capture the impact of variability in these parameters. Evidence suggests that net use fluctuates annually [44], and that exposure to infectious bites may be equally variable given human and mosquito activity [45]. Parameterising an individual-based model with field estimates of this inter and intra season nightly variability and within-cluster variability could substantially reduce the discrepancy between observed data and model predictions.

Our analysis indicates that the mass community effect is powerful, offering substantial protection to both those using and not using mosquito nets within the community. Proving this using empirical measures has many challenges, though a recent systematic review came to similar conclusions. The best evidence verifying this result comes from early trials in Ghana and Kenya testing the utility of insecticide treated nets (dip-nets that could be re-treated every 6 months) which indicated epidemiological protection in residents of the ITN-treated villages, and additionally for households without ITNs that were up to a few hundred metres away [15, 46]. Our models show the magnitude of the community effect is highly variable and will change according to levels of endemicity, LLIN use and factors such as insecticide resistance. It is also likely to vary with the fine-scale geography of the communities (how houses and LLINs are clustered) and the local flying and egg-laying behaviour of the local mosquito vectors. Given this variability it is unsurprising that observational studies vary substantially.

The modelling framework allows us to untangle the community protection offered from the barrier and insecticide, which stresses the additional benefit elicited from killing mosquitoes. Importantly we parameterise the model with results from experimental hut trials evaluating both insecticide treated and untreated nets as the physical barrier of the mosquito net has been shown to induce some mortality in mosquitoes as they attempt to feed [32]. Nevertheless, the relative protection provided by the insecticide is considerable, the reduction in EIR for LLINs is 89% [95% CI: 67% - 98%] for users and 74% [95% CI: 48% - 92%] for non-users in areas with an EIR of 100 prior to the introduction of 80% net use. In contrast, the total protection offered by an untreated net used by 80% of the population is 52% [95% CI: 12% - 84%] for users and 21% [95% CI: 0% - 57%] for non-users

Results illustrate the importance of achieving high coverage with both LLINs and untreated nets as increasing net use always increases protection. In the setting examined here the benefit provided to a user increase by 27% if they are in a community where 80% of people use LLINs. Interestingly, the model predicts that the direct benefit of the barrier and the insecticide diminishes in users as usage increases in the community. The phenomenon can be explained by more widespread net use means that net users are more likely to experience biting attempts made by mosquitoes that have previously tried to feed on other net users. This decrease in the direct protection of the barrier is more than compensated for by the indirect protection provided by the barrier and the insecticide which substantially increases as net use increases. The benefit of the community effect is observed across the range of scenarios examined indicating that there is no minimum LLIN usage needed for users and non-users to benefit from community protection.

The analysis explores what happens as an increasing proportion of local mosquitoes can survive exposure to pyrethroid insecticide. It is predicted that the diminished killing depletes the community effect from LLINs for both net users and non-users (Figure 4). The emergence of pyrethroid resistance in *Anopheles* mosquitoes, alongside a recognition for local communities to lead intervention provision and plug gaps in availability of LLINs, has opened up a conversation on the necessity for insecticide in nets [10, 47]. Here, we can see that from an epidemiological perspective the capacity to kill mosquitoes is critical for the protective impact of these net interventions. The work suggests that maximising the proportion of people covered by LLINs should always be the aim but then any additional overall increase of net use with untreated nets will also be beneficial. Novel net designs that either chemically [48–50] or mechanically [51] kill mosquitoes are welcome to mitigate for the potential loss in impact from pyrethroid-LLINs [35, 52].

There was no measurable difference between disease prevalence in users and non-users in the DHS data over time and for our estimates of the level of pyrethroid resistance in the local mosquito population. Previous studies have suggested that higher infection rates in non-users than users seen in recent data may indicate reduced community protection [39]. Our analysis of DHS data does not support this suggestion as there is no evidence that the difference between users has changed over the time-period during which resistance has spread. The analytical work provides a reason for why this might be the case as though the model predicts a reduction in the difference between users and non-users as pyrethroid resistance increases (Figure S2) the magnitude of this change is relatively modest in relation to the differences caused by the barrier effect of the net. Care should be taken interpreting the DHS data as our estimates of the level of resistance in a community using the discriminating dose bioassay are highly variable and have been shown to vary substantially at the district level. The level of resistance in each DHS cluster was not directly measured but instead inferred, reducing the power of the analyses, and ultimately making it under-powered to detect the differences suggested by the model. Thomas and Read [53] suggest that it is the indirect, or community, protection that is at greatest risk with increasing pyrethroid resistance for countries with moderate-to-high prevalence but incomplete ITN coverage. Our modelling results further suggest that it is both the direct and indirect impact of the insecticide that are impacted by pyrethroid resistance, but the impact on the indirect effect is larger (Figure 4, Figure S6, Figure S7 and Figure S8).

The principal limitations in our analysis are related to the modelling approach. We do not parameterise the transmission model to match the specific ecology of each DHS cluster, opting instead (given restrictions in data) only to calibrate to the respective endemicity reported across the communities in the DHS data. It is therefore reassuring that we broadly reflect the trends in the user-non-user differences in malaria burden across usage and endemicity levels, but not surprising that we do not capture the magnitude given the reasons stated above. There may be subtle differences in the performance of net products [38, 54], but we assume all pyrethroid LLINs to have equivalent impact regardless of brand. Our assumptions are also fixed throughout the year whereas there are certain to be differences in individual daily use, risk of bites given weather conditions and seasonal mosquito abundance and the access to treatment of different communities represented in the DHS data among other factors. It should be stressed that these modelling results are dependent on the assumptions made in the modelling process (for example, what happens to a mosquito when they are dissuaded from feeding on someone who is using a net, outlined in full in the supplement). Though these assumptions reflect our current best understanding of the processes involved, they should be verified, for example using recently developed technology that can track the mosquito in the home [55].

Factors governing the use of LLINs in communities at risk of malaria are complex [4]. This work indicates that increasing LLIN usage is always beneficial to control malaria, through both direct and indirect protection. Maximum direct protection from indoor interventions is limited in places with higher levels of outdoor biting (be it biting during the early evening, later morning or daytime) even if no pyrethroid resistance exists and everyone uses LLINs [56]. The indirect protection provided by the insecticide can help overcome the issues of residual transmission though this benefit is diminished as mosquitoes develop resistance to the insecticide [56, 57]. The importance of the killing effect of insecticidal nets cannot be under-estimated because it causes a substantial reduction in the number of infectious bites received per person per year, proving a crucial part of the mass community effect and contributing hugely to the success associated with LLIN interventions [1, 58, 59]. Determining the different protective effects of LLINs allows us to better understand how insecticide resistance is likely to impact on malaria control, and how best to mitigate against it. Appreciating all aspects of protection that are afforded by LLINs will be crucial in the drive for malaria control – helping to shape how we can innovate nets with improved effectiveness – particularly in the reality of insecticide resistance.

## Data Availability

The deterministic malaria model used for this analysis can be found on GitHub (https://github.com/mrc-ide/dide-deterministic-malaria-model). The Demographic Health Survey data is publicly available at https://dhsprogram.com.

https://github.com/mrc-ide/dide-deterministic-malaria-model

## Acknowledgements

HJTU was funded by a grant from the Gates foundation and an Imperial College Research Fellowship. ESS is funded by a UKRI Future Leaders Fellowship from the Medical Research Council (MR/T041986/1). Funding support for TSC and ESS was also received from the Innovative Vector Control Consortium through the New Nets Project funded by Unitaid, the Wellcome Trust [200222/Z/15/Z] MiRA. All authors acknowledge funding from the MRC Centre for Global Infectious Disease Analysis (reference MR/R015600/1), jointly funded by the UK Medical Research Council (MRC) and the UK Foreign, Commonwealth & Development Office (FCDO), under the MRC/FCDO Concordat agreement and is also part of the EDCTP2 programme supported by the European Union; and acknowledges funding by Community Jameel.

## Supplementary Information

**S1 Text. Model description**

**Table S1.** Uncertainty parameter ranges used to explore uncertainty. The untreated and treated net (LLIN) parameters are shown for specified levels of resistance where 0 indicates the absence of pyrethroid resistance (100% of mosquitoes are killed on exposure to a discriminatory dose of pyrethroid insecticide during bioassay testing), 40 and 80 indicate 60% and 20% of mosquitoes are killed on exposure respectively during discriminatory dose bioassay testing.

| Parameter | Description | Value | Reference |
| --- | --- | --- | --- |
| $\Phi_B$ | Probability of a mosquito bite in bed in the absence of nets | 0.85 (0.53 – 0.98) | Sherrard-Smith et al 2019 [56] |
| Untreated nets |  |  |  |
| $d_{NO}$ | Net efficacy parameter determining the probability of dying | 0.059 (0.012-0.125) | This paper, derived from Nash [32] |
| $r_{NO}$ | Net efficacy parameter determining the probability of successfully feeding | 0.409 (0.198-0.538) | |
| $\gamma_N$ | Net efficacy parameter determining half-life (which is included for untreated nets to match the durability of LLINs to reflect an assumption of wear over time). | 2.64 | |
| Treated nets (LLINs) |  |  |  |
| $d_{NO}$ | Net efficacy parameter determining the probability of dying | 0.387 (0.314 – 0.451) | Sherrard-Smith et al in review |
| $r_{NO}$ | Net efficacy parameter determining the probability of repeating | 0.563 (0.626 – 0.513) | |
| $\gamma_N$ | Net efficacy parameter determining half-life (to reflect the waning insecticidal impact and wear over time) | 2.64 (2.000 – 3.000) years | |

**Table S2:** DHS Surveys used in analysis.

| **Country** | **Code** | **Years** |
| --- | --- | --- |
| Angola | AO | 2015 |
| Benin | BJ | 2012, 2017 |
| Burkina Faso | BF | 2010, 2014, 2017 |
| Burundi | BU | 2012 |
| DRC | CD | 2013 |
| Cote d'Ivoire | CI | 2012 |
| Ghana | GH | 2014, 2016, 2019 |
| Guinea | GN | 2012 |
| Kenya | KE | 2015 |
| Liberia | LB | 2011, 2016 |
| Madagascar | MD | 2011, 2013, 2016 |
| Malawi | MW | 2012, 2014, 2017 |
| Mali | ML | 2012, 2015, 2018 |
| Mozambique | MZ | 2011, 2015, 2018 |
| Nigeria | NG | 2010, 2015, 2018 |
| Rwanda | RW | 2010, 2015 |
| Senegal | SN | 2010, 2012, 2014, 2015, 2016 |
| Sierra Leone | SL | 2016 |
| Tanzania | TZ | 2012, 2015, 2017 |
| Togo | TG | 2013, 2017 |
| Uganda | UG | 2014, 2016, 2018 |

**Figure S1.**
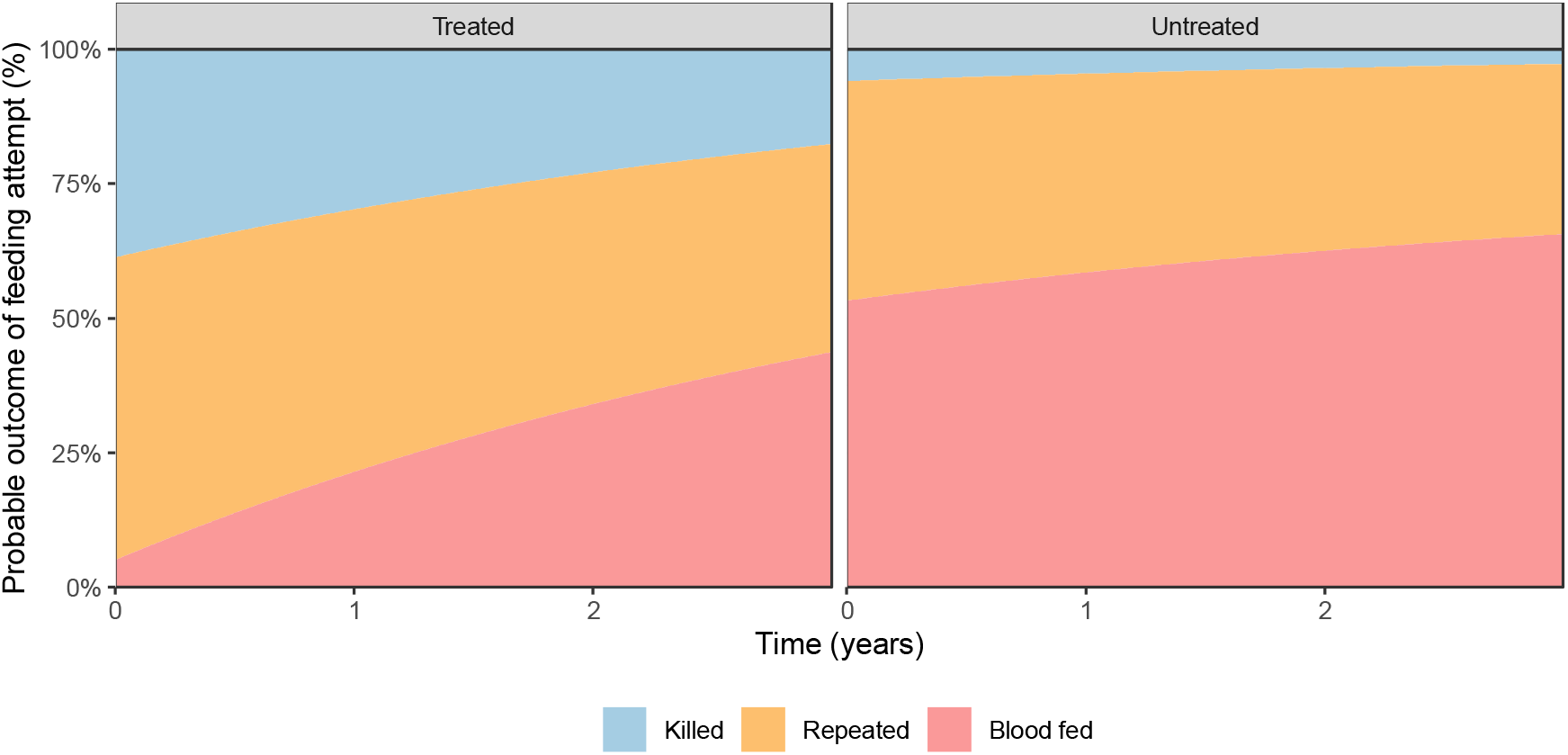
Changes in probable outcomes of feeding attempts for Treated and Untreated nets. The red area shows the percentage of mosquitoes that will die, green that will repeat and blue that will successfully feed. Models were fit to data summarised by Nash et al. using the parameter estimates given in Table S1.

**Figure S2.**
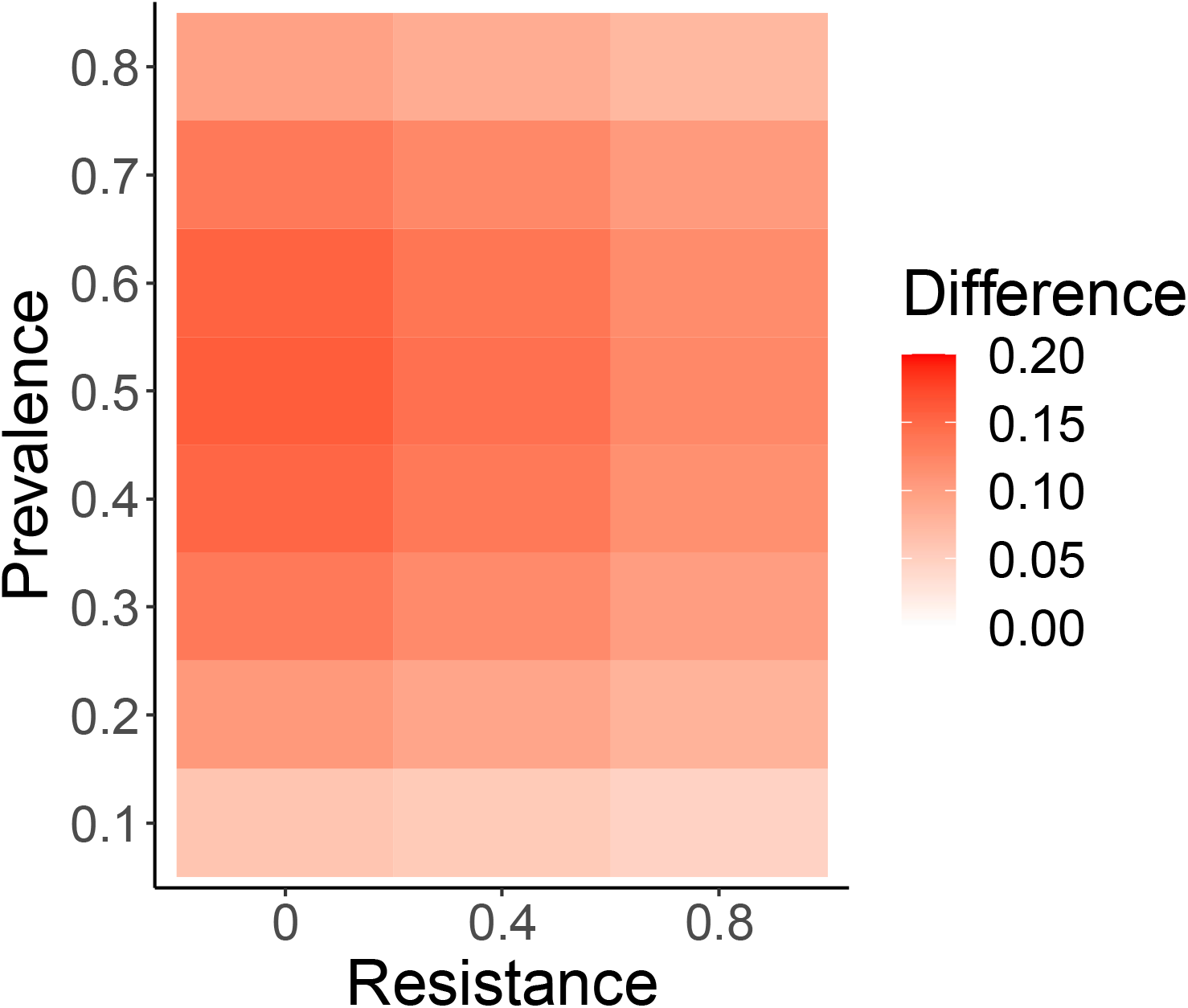
Transmission model estimates of the absolute difference in prevalence between users and non-users of LLINs for varying endemicities of disease (slide prevalence) and levels of pyrethroid resistance in the local mosquito population. An overall LLIN usage of 50% is assumed throughout. The level of resistance assumed by the model is measured as the percentage of mosquitoes surviving a discriminating dose bioassay.

**Figure S3.**
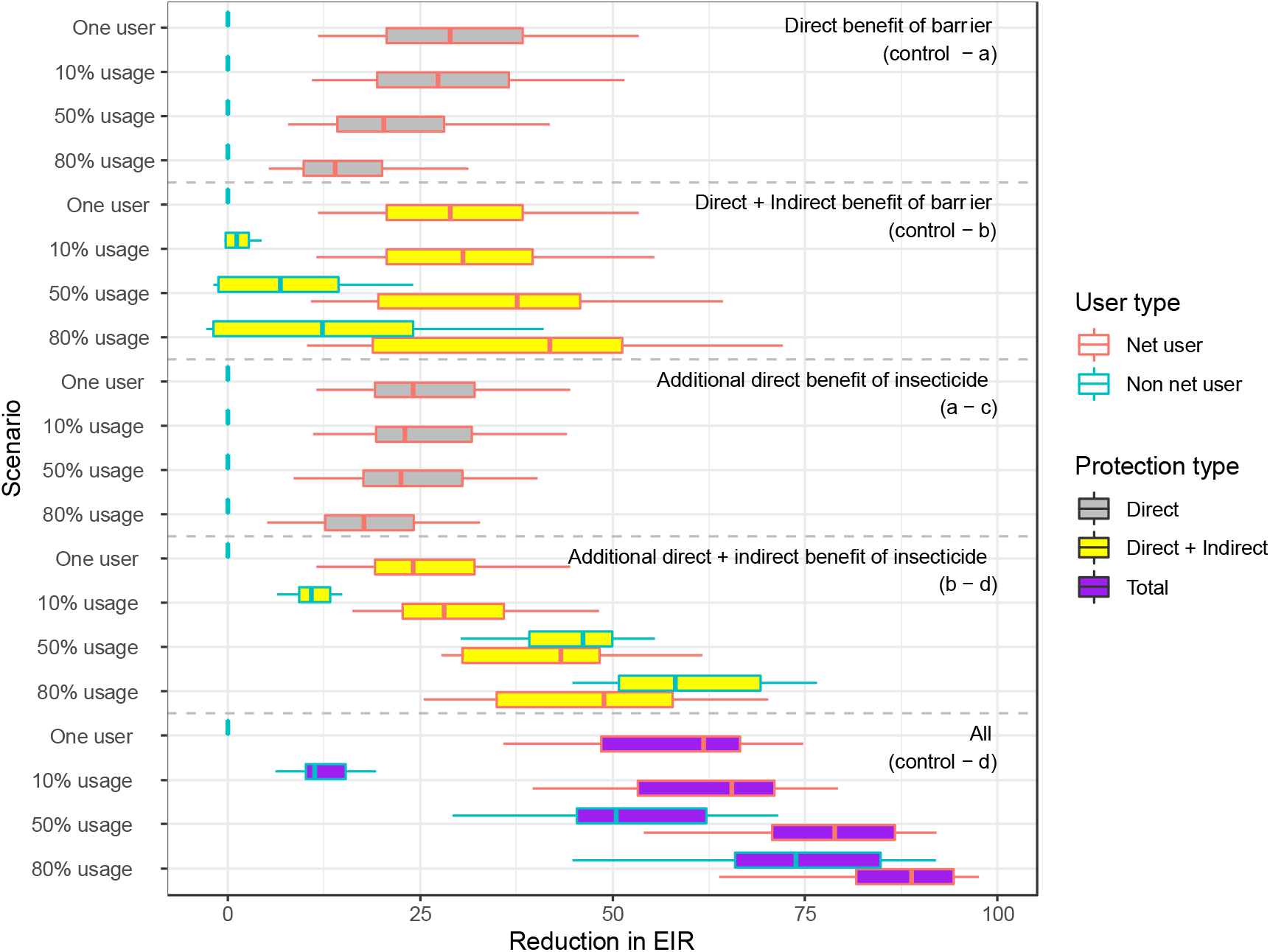
Reduction in EIR from direct and direct + mass community (indirect) protection offered by mosquito nets for a pre-intervention EIR of 100. The reduction in EIR is calculated from a control, where nobody in the population is given a net, for the 5 scenarios detailed in the methods section. Scenarios (a) – (d) are repeated for an individual using a net and 10%, 50% and 80% of the population using nets. The reduction for users is shown in red and for non-users is shown in blue. Direct reductions in EIR are filled in grey, indirect reductions in yellow and total reductions in purple. The total protection offered by an untreated net is shown here as the direct + indirect benefit of barrier.

**Figure S4:**
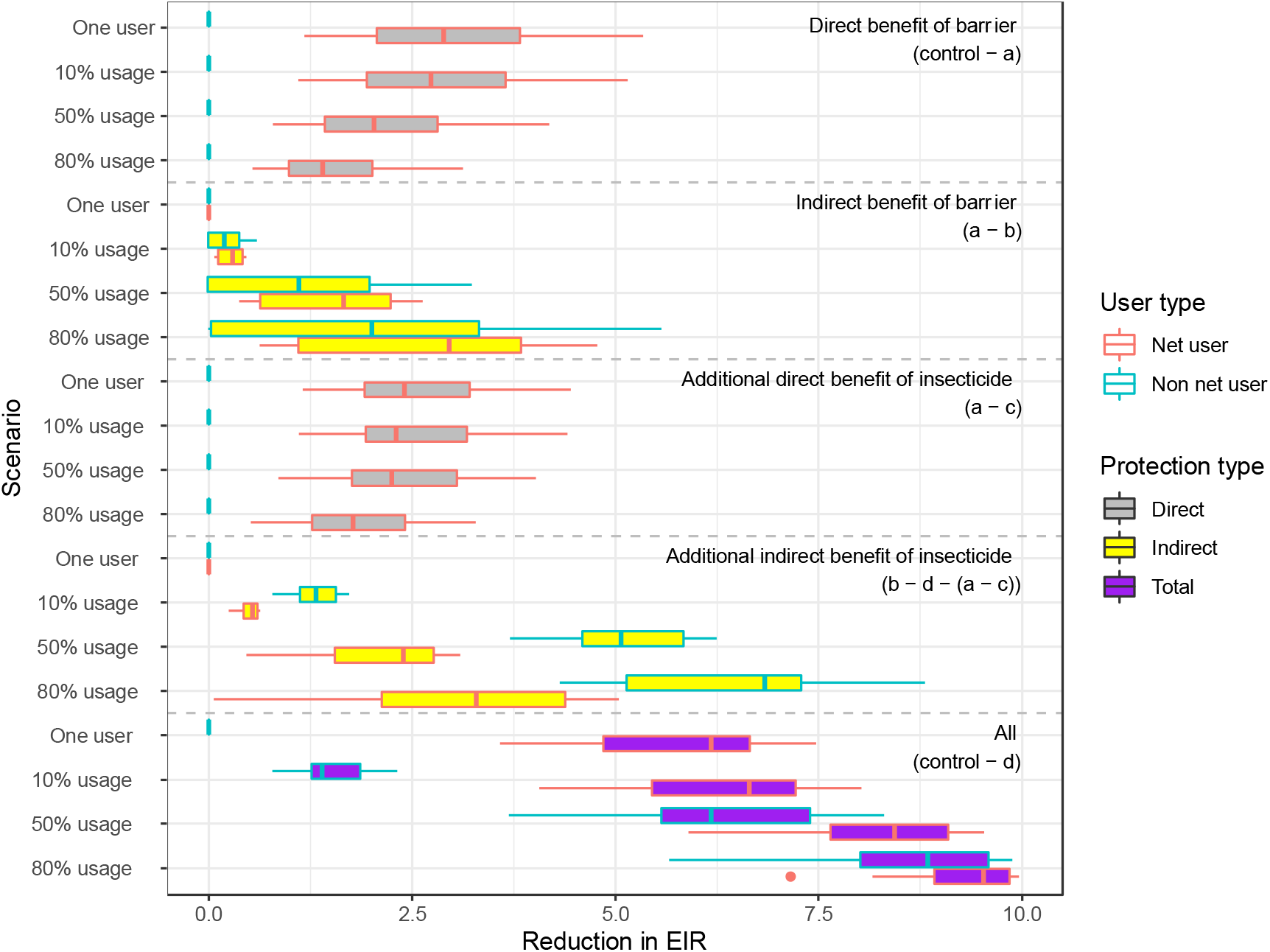
Reduction in EIR from direct and mass community (indirect) protection offered by mosquito nets for a pre-intervention EIR of 10. The reduction in EIR is calculated relative to a control scenario, where nobody in the population is given a net, for the 8 scenarios detailed in the methods section. Scenarios (a) – (d) are repeated for an individual using a net and 10%, 50% and 80% of the population using nets. The reduction for users is shown in red and for non-users is shown in blue. Direct reductions in EIR are filled in grey, indirect reductions in yellow and total reductions in purple.

**Figure S5:**
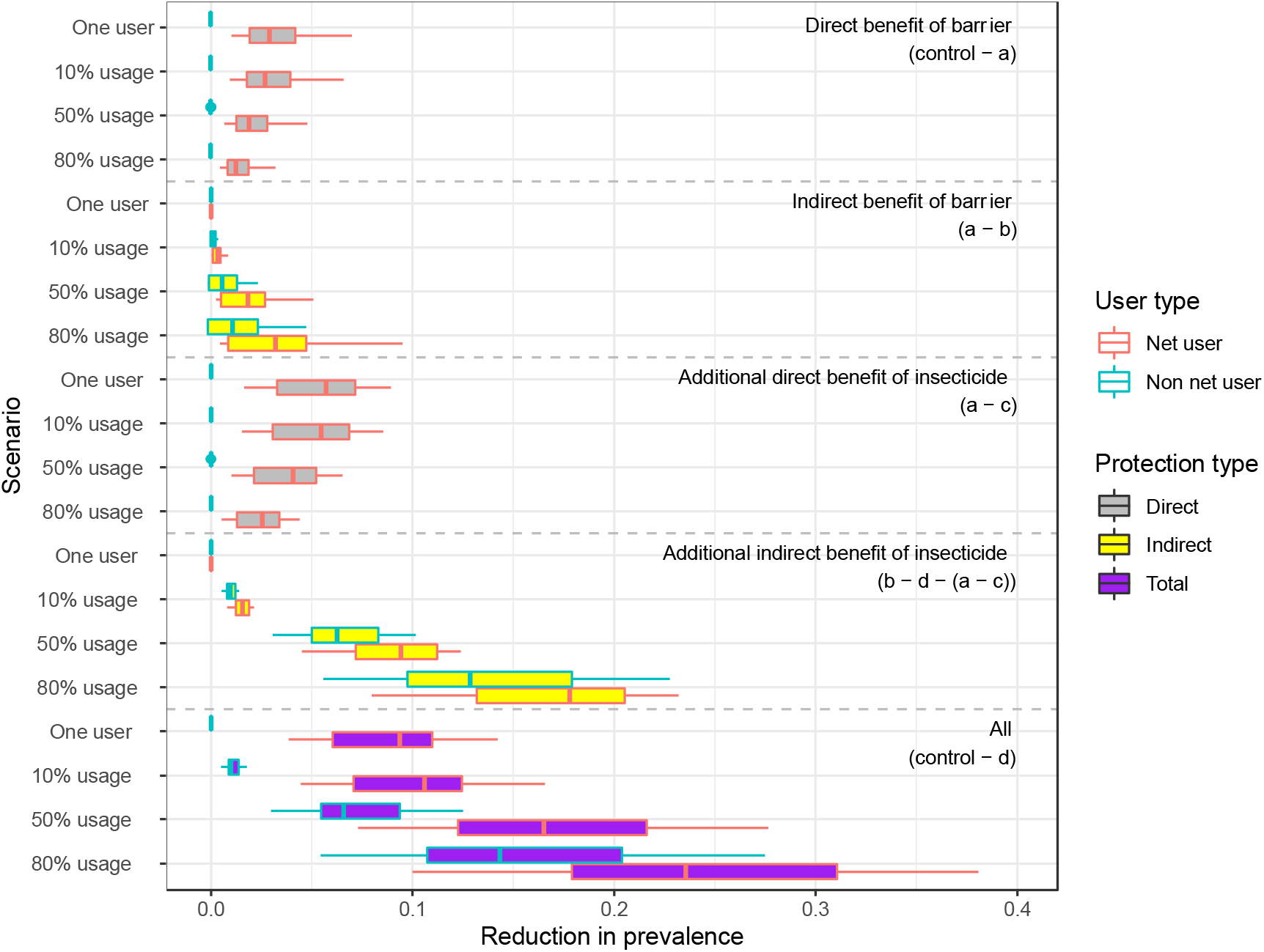
Reduction in prevalence from direct and mass community (indirect) protection offered by mosquito nets for a pre-intervention EIR of 100. The reduction in prevalence is calculated from a control, where nobody in the population is given a net, for the 8 scenarios detailed in the methods section. Scenarios (a) – (d) are repeated for an individual using a net and for 10%, 50% and 80% of the population using nets. The reduction for users is shown in red and for non-users is shown in blue. Direct reductions in EIR are filled in grey, indirect reductions in yellow and total reductions in purple.

**Figure S6:**
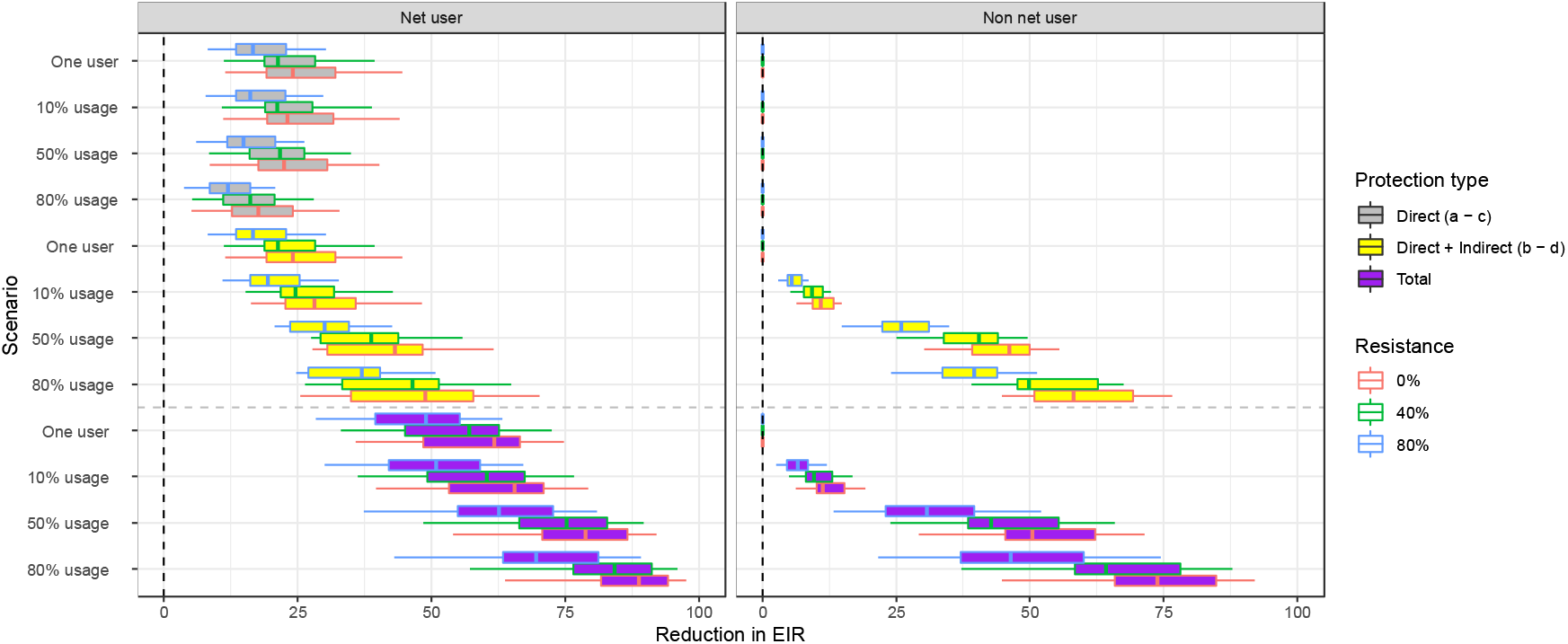
Reduction in EIR from direct and direct + indirect protection offered by LLIN at differing levels of pyrethroid resistance for a pre-intervention EIR of 100. Left figure shows the effect for users and right figure shows the effect for non-users. The reduction in EIR is calculated from a control, where nobody in the population is given a net, for the scenarios detailed in the methods section where insecticide is included. Usages of 1 person in the population, 10%, 50% and 80% are considered for 0% resistance (as in *S1* Figure) and 40% and 80% resistance. The barrier only effect remains constant at differing levels of resistance so are not included.

**Figure S7:**
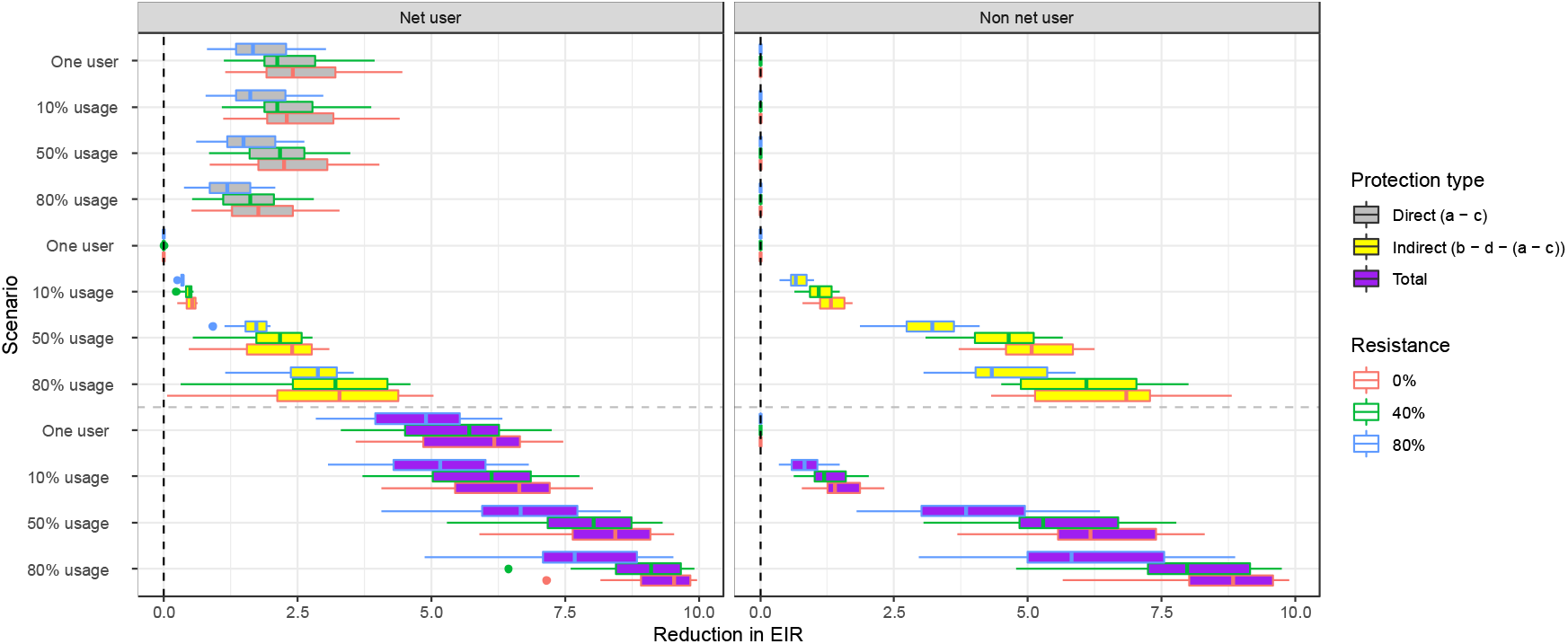
Reduction in EIR from mass community protection offered by LLIN at differing levels of pyrethroid resistance for a pre-intervention EIR of 10. Left figure shows the effect for users and right figure shows the effect for non-users. The reduction in EIR is calculated from a control, where nobody in the population is given a net, for the scenarios detailed in the methods section where insecticide is included. Usages of 1 person in the population, 10%, 50% and 80% are considered for 0% resistance (as in Figure 3) and 40% and 80% resistance. The barrier only effect remains constant at differing levels of resistance so are not included.

**Figure S8:**
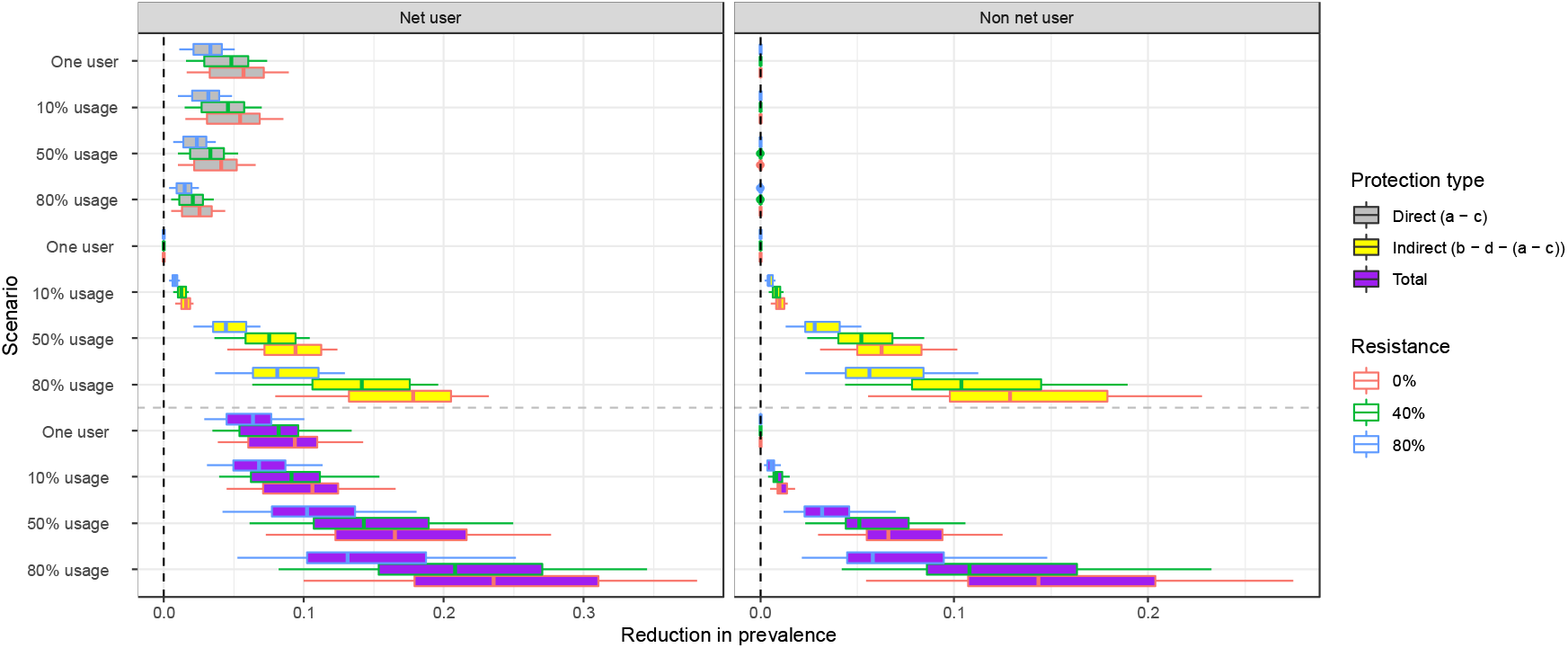
Reduction in prevalence from direct and mass community (indirect) protection offered by LLIN at differing levels of pyrethroid resistance for a pre-intervention EIR of 100. Left figure shows the effect for users and right figure shows the effect for non-users. The reduction in prevalence is calculated from a control, where nobody in the population is given a net, for the scenarios detailed in the methods section where insecticide is included. Usages of 1 person in the population, 10%, 50% and 80% are considered for 0% resistance (as in Figure S5) and 40% and 80% resistance. The barrier only effect remains constant at differing levels of resistance so are not included.

