## Supplementary Text 1 for "Quantifying the direct and indirect protection provided by insecticide treated bed nets against malaria"

### Modelling the impact of pyrethroid resistance on the mass community effect of insecticide treated nets

H. Juliette T. Unwin[1], Ellie Sherrard-Smith[1], Thomas S. Churcher[1], Azra C. Ghani[1]
[1]*MRC Centre for Global Infectious Disease Analysis, Department of Infectious Disease Epidemiology, Faculty of Medicine, Imperial College London*

#### Supplementary Information 1: Imperial deterministic malaria model

We use a previously published malaria transmission model that fully incorporates the dynamics of *Plasmodium falciparum* transmission between human and vector hosts. The model is deterministic and similar to that presented in [1] and in [2].

##### The human model

Our model groups people within a population into compartments based on their age, which are denoted by the subscript $i$. At each point in time the age compartments can be in one of six infectious states – susceptible ( $S_i$ ), treated clinical disease ( $T_i$ ), untreated clinical disease ( $D_i$ ), asymptomatic patent infection ( $A_i$ ), sub-patent infection ( $U_i$ ) and protected by a period of prophylaxis from prior treatment ( $P_i$ ). People, or proportions of the population in each compartment, move between these states as shown in Figure A with the rates marked on the arrows and described below.

People are born into the first age compartment of this model as susceptible to infection, with new-borns possessing a level of maternally-inherited immunity that decays over the first six months of their lives. They move through the age compartments as expected due to natural aging. A proportion of each susceptible compartment is exposed to infectious bites from the mosquito vector as time passes. The hazard of infection for each compartment is determined by the force of infection $(\Lambda_i)$, which is a function of the compartmental pre-erythrocytic immunity and biting rate, and the mosquito population size and level of infectivity.

Proportions of the infected compartment develop clinical disease or asymptomatic infection following a latent period $(\tau_E)$ (and move to compartments D or A) depending on the probability of acquiring clinical disease $(\phi_i)$, which is dependent on the compartments level of clinical immunity and is defined in equation (9). The proportion who develop clinical disease can be successfully treated (with a fixed probability $f_T$ ) and move to infection state *T*, or will not seek treatment (with probability $1-f_T$ ) and move to infection state *D*. Proportions of the treated compartment then recover from infection at rate $r_T$ and return to the susceptible infection state *S*. However, they retain a degree of drug-dependent partial protection from reinfection (modelled as a Weibull survivorship curve) which wanes over time [3].

The proportion of the untreated clinical disease state (D) compartment not receiving treatment recover to the asymptomatic infection state (*A*) at rate $r_D$ . Death is not explicitly modelled. As parasite density is controlled, proportions of the asymptomatic state compartment progress to the

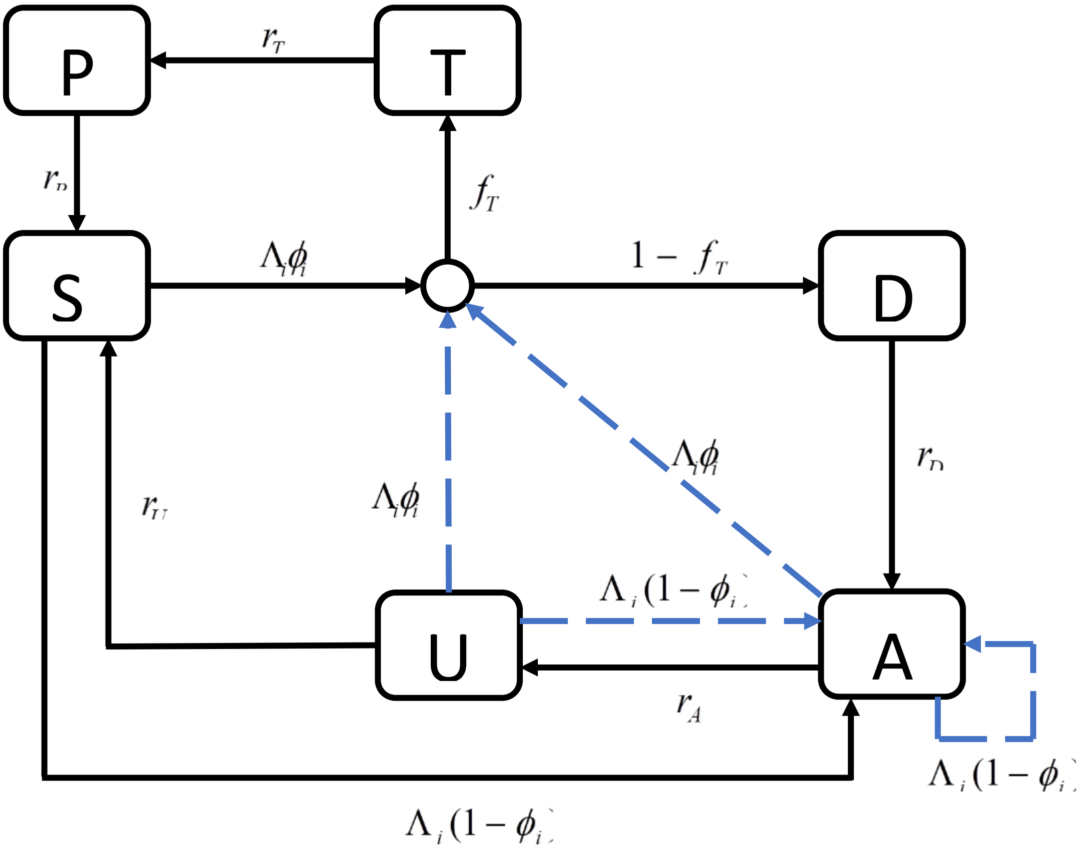


**Figure A: Illustration of the progression between human infection states** (S = susceptible, D = clinical disease, T = successfully treated disease, A = asymptomatic patent infection, U = asymptomatic subpatent infection). The states are shown in boxes with the transitions marked by arrows and associated hazard rates. The circle represents the treatment node. The dashed blue arrows indication superinfection.

sub-patent infection state (*U*) with rate $r_A$ , before naturally clearing infection and returning to susceptible (*S*) with rate $r_U$ .

Superinfection is included with all age and heterogeneity compartments in states *D*, *A* and *U.* This means that these compartments remain susceptible to re-infection. If this occurs, the proportions of the compartments move into infection states *D*, *T* or *A* in the same process described above. All rates are constant and independent of age.

This model is described by the following coupled ordinary differential equations:

$$\begin{aligned}
\frac{\partial S_i(t)}{\partial t}+\frac{\partial S_i(t)}{\partial a} &= -\Lambda_i(t-d_E)S_i(t)+r_pP_i(t)+r_uU_i(t),\\
\frac{\partial T_i(t)}{\partial t}+\frac{\partial T_i(t)}{\partial a} &= \phi_i f_T\Lambda_i(t-d_E)(S_i(t)+A_i(t)+U_i(t))-r_TT_i(t),\\
\frac{\partial D_i(t)}{\partial t}+\frac{\partial D_i(t)}{\partial a} &= \phi_i(1-f_T)\Lambda_i(t-d_E)(S_i(t)+A_i(t)+U_i(t))-r_DD_i(t),\\
\frac{\partial A_i(t)}{\partial t}+\frac{\partial A_i(t)}{\partial a} &= (1-\phi_i)\Lambda_i(t-d_E)(S_i(t)+U_i(t))+r_DD_i(t)-\phi_i\Lambda_i(t-d_E)A_i(t)-r_AA_i(t),\\
\frac{\partial U_i(t)}{\partial t}+\frac{\partial U_i(t)}{\partial a} &= r_AA_i(t)-r_UU_i(t)-\Lambda_i(t-d_E)U_i(t),\\
\frac{\partial P_i(t)}{\partial t}+\frac{\partial P_i(t)}{\partial a} &= r_TT_i(t)-r_PP_i(t).
\end{aligned} \qquad (1)$$

The parameters for equation (1) are given in Table S1.1. Equations for $\Lambda_i$ and $\phi_i$ are evaluated below in equations (4) and (9) respectively.

**Table S1.1: The human model parameters**

| Parameter | Symbol | Estimate |
|---|---|---|
| Latent period | $\tau_E$ | 12 days |
| **Rate of leaving human infection stages** | | |
| Asymptomatic patent infection | $r_A$ | 0.00512821 |
| Treated clinical disease | $r_T$ | 0.2 |
| Untreated clinical disease | $r_D$ | 0.2 |
| Asymptomatic patent infection | $r_U$ | 0.00906627 |
| Prophylaxis | $r_P$ | 0.06666667 |
| **Treatment Parameters** | | |
| Probability of seeking treatment if clinically diseased | $f_T$ | 0.4 |

#### Heterogeneity in biting rates

Each age compartment is assigned a unique biting rate, $\psi_i$. This is defined as:

$$\psi_i(a)=1-\rho\exp\left(-\frac{a_i}{a_0}\right), \qquad (2)$$

for age compartment $a_i$, where $\rho$ and $a_0$ are parameters that determine the relationship between age (i.e. body size) and biting rate. The relative biting rate, $\zeta_j$, is drawn from a log-normal distribution with a mean of 1:

$$\log(\zeta_j)\sim N\left(\frac{-\sigma^2}{2},\sigma^2\right), \qquad (3)$$

where $j$ denotes the heterogeneity group of the model.

The entomological inoculation rate (EIR), $\varepsilon_{i,j}$ (or the average number of infecting bites an individual receives), and force of infection, $\Lambda_{i,j}$, experienced by an age compartment $i$ and heterogeneity compartment $j$, with age $a$ at time $t$ are denoted as:

$$\begin{aligned}\varepsilon_{i,j} &= \varepsilon_0(t)\zeta_j\psi_i(a),\\ \Lambda_{i,j} &= b_i(t)\varepsilon_{i,j}(a,t),\end{aligned} \tag{4}$$

where $\varepsilon_0(t)$ is the mean EIR experienced by adults at time $t$, and $b_i(t)$ is the probability that an infectious bite leads to a patent infection, which is determined by the level of pre-erythrocytic immunity and is defined in equation (5). The force of infection is then subject to a lag of $\tau_E$ days to account for the latent period of infection.

The parameters for this section are given in **Table S1.*2***. Descriptions for $b_i$ and $\varepsilon_0$ are given in equations (6) and (23).

**Table S1.2: Age and heterogeneity parameters**

| Parameter | Symbol | Estimate |
|---|---|---|
| Age-dependent biting parameter | $\rho$ | 0.85 |
| Age-dependent biting parameter | $a_0$ | 8 years |
| Variance of the log heterogeneity in biting rates | $\sigma^2$ | 1.67 |

#### Human immunity

The acquisition and loss of naturally-acquired immunity is captured dynamically in the model and is driven by both age and exposure. We consider three transition points at which immunity may act:

1) a reduction in the probability that infection is established following an infectious challenge (pre-erythrocytic immunity, $I_B$),
2) a reduction in the probability of clinical disease upon infection (clinical immunity, $I_C$),
3) and a reduction in the detectability of an infection and onward transmission to mosquitoes through blood stage immunity (detection immunity, $I_D$).

Immunity to infection, $I_{B_{i,j}}(t)$ in a population exposed to an EIR $\varepsilon_{i,j}(t)$ is a function of both age and time and is given by the partial differential equation,

$$\frac{\partial I_{B_{i,j}}}{\partial t} + \frac{\partial I_{B_{i,j}}}{\partial a} = \frac{\varepsilon_{i.j}}{\varepsilon_{i.j}u_B + 1} - \frac{I_{B_{i,j}}}{d_B}, \qquad I_{B_{i,j}}(0,t) = 0 \tag{5}$$

where $u_B$ limits the rate at which immunity to infection can be boosted at high exposure and $d_B$ is the mean duration of immunity to infection. The probability of infection by age is then given by a Hill function,

$$b_{i,j}(t) = b_0\left(b_1 + \frac{1-b_1}{1+\left(\frac{I_{B_{i,j}}(t)}{\mathrm{I}_{B0}}\right)^{\kappa_B}}\right), \tag{6}$$

where $b_0$ is the probability of infection with no immunity, $b_0 b_1$ is the minimum probability, $I_{B0}$ and $\kappa_B$ are scale and shape parameters respectively, and $I_{B_{i,j}}(t)$ is the level of pre-erythrocytic immunity of age compartment $i$ and heterogeneity compartment $j$ at time $t$.

Immunity to clinical disease, $I_{C_{i,j}}(t)$, comprises of immunity acquired by exposure to infection, $I_{CA_{i,j}}$, and that maternally acquired, $I_{CM_{i,j}}$. $I_{CA_{i,j}}(t)$ in a population exposed to a force of infection $\Lambda_{i,j}$ is a function of both age, heterogeneity group and time and is described by the partial differential equation:

$$\frac{\partial I_{CA_{i,j}}}{\partial t} + \frac{\partial I_{CA_{i,j}}}{\partial a} = \frac{\Lambda_{i,j}}{\Lambda_{i,j}\, u_C + 1} - \frac{I_{CA_{i,j}}}{d_{CA}}, \qquad I_{CA_{i,j}}(t) = 0 \tag{7}$$

where $u_C$ limits the rate at which immunity to clinical disease can be boosted at high exposure and $d_{CA}$ is the mean duration of clinical immunity. Maternally-acquired immunity $I_{CM_{i,j}}(t)$ is assumed at birth to be a proportion ($P_{CM}$) of the level of immunity present in a 20-year old woman, $I_{C_{20,j}}(t)$, living in the same location and which decays at a constant rate ($1/d_M$),

$$\frac{\partial I_{CM_{i,j}}}{\partial t} + \frac{\partial I_{CM_{i,j}}}{\partial a} = -\frac{I_{CM_{i,j}}}{d_M}, \qquad I_{CM_{i,j}}(t) = P_{CM} I_{C_{20,j}}(t). \tag{8}$$

The total clinical immunity by age and time is given by $I_C = I_{CA} + I_{CM}$. The probability of acquiring clinical disease upon infection by age is then given by a Hill function,

$$\phi_{i,j}(t) = \phi_0\left(\phi_1 + \frac{1-\phi_1}{1+\left(\frac{(I_{CA_{i,j}}(t) + I_{CM_{i,j}}(t)}{I_{C0}}\right)^{\kappa_C}}\right) \tag{9}$$

where $\phi_0$ is the probability of disease with no immunity, $\phi_0\phi_1$ is the minimum probability, $I_{C0}$ and $\kappa_C$ are scale and shape parameters respectively, $I_{CA_{i,j}}(t)$ is the level of acquired immunity to clinical disease and $I_{CM_{i,j}}(t)$ is the level of maternally acquired immunity to clinical disease of age compartment $i$ and heterogeneity class $j$ at time $t$.

Finally, detection immunity, $I_{D_{i,j}}$, which is the effect of blood stage immunity reducing the detectability of an infection and onward transmission to mosquitoes, is given by the partial differential equation:

$$\frac{\partial I_{D_{i,j}}}{\partial t}+\frac{\partial I_{D_{i,j}}}{\partial a}=\frac{\Lambda_{i,j}}{\Lambda_{i,j}u_D+1}-\frac{I_{D_{i,j}}}{d_D}, \qquad I_{D_{i,j}}(t)=0, \tag{10}$$

where $u_D$ limits the rate at which detection immunity can be boosted at high exposure and $d_C$ is the mean duration of detection immunity. The detectability by microscopy of an asymptomatic infection in age compartment $i$ and heterogeneity compartment $j$ at time $t$ is given by:

$$q_{i,j}(t)=d_1+\frac{(1-d_1)}{\left(\left(\frac{1+I_{D_{i,j}}(t)}{I_{D0}}\right)^{\kappa_D} f_{D_i}\right)} \tag{11}$$

where $d_1$ is the minimum probability of detection, $I_{D0}$ and $\kappa_D$ are scale and shape parameters respectively, $I_{D_{i,j}}(t)$ is the level of acquired immunity to the detectability of infection of age compartment $i$ and heterogeneity compartment $j$ at time $t$, and

$$f_{D_i}=1-\frac{(1-f_{D0})}{\left(1+\left(\frac{a_i}{a_D}\right)^{\gamma_D}\right)} \tag{12}$$

is an age-dependent (physiological) modifier of the detectability of infection where $f_{D0}$, $a_D$ and $\gamma_D$ are parameters.

The immunity parameters are given in Table S1.3.

**Table S1.3: Immunity parameters**

| Parameter | Symbol | Estimate |
|---|---|---|
| **Immunity reducing probability of infection** | | |
| Maximum probability due to no immunity | $b_0$ | 0.590076 |
| Maximum relative reduction due to immunity | $b_1$ | 0.5 |
| Inverse of decay rate | $d_B$ | 10 years |
| Scale parameter | $I_{B0}$ | 43.8787 |
| Shape parameter | $\kappa_B$ | 2.15506 |
| Duration in which immunity is not boosted | $u_B$ | 7.19919 days |
| **Immunity reducing probability of clinical disease** | | |
| Maximum probability due to no immunity | $\phi_0$ | 0.791666 |
| Maximum relative reduction due to immunity | $\phi_1$ | 0.000737 |
| Inverse of decay rate | $d_{CA}$ | 30 years |
| Scale parameter | $I_{C0}$ | 18.02366 |
| Shape parameter | $\kappa_C$ | 2.36949 |
| Duration in which immunity is not boosted | $u_C$ | 6.06349 days |
| Inverse of decay rate of maternal immunity | $d_M$ | 67.6952 days |
| New-born immunity relative to mother's | $P_{CM}$ | 0.774368 |
| **Immunity reducing probability of detection** | | |
| Minimum probability due to maximum immunity | $d_1$ | 0.160527 |
| Inverse of decay rate | $d_{ID}$ | 10 years |
| Scale parameter | $I_{D0}$ | 1.577533 |
| Shape parameter | $\kappa_D$ | 0.476614 |
| Duration in which immunity is not boosted | $u_D$ | 9.44512 days |
| Scale parameter relating age to immunity | $a_D$ | 21.92 years |
| Time-scale at which immunity changes with age | $f_{D0}$ | 0.007055 |
| Shape parameter relating age to immunity | $\gamma_D$ | 4.8183 |

#### Vector model

The larval model is based on the compartmental model previously described in [4]. Female adult mosquitoes $(M)$ lay eggs at rate $\beta_L$. Upon hatching from eggs, larvae progress through early and late larvae stages ($E_L$ and $L_L$ compartments) before developing into the pupal stage $(P_L)$. The duration spent in each stage is denoted by $d_{E_L}$, $d_{L_L}$ and $d_{P_L}$ respectively. The larval stages are regulated by density dependent mortalities ($\mu_{E_L}$, $\mu_{L_L}$ and $\mu_{P_L}$) with a time-varying carrying-capacity, $K_L$, which represents the ability of the environment to sustain breeding sites through different periods of the year, and with the density of larvae in relation to the carrying-capacity regulated by a parameter $\gamma_L$. The carrying-capacity determines the mosquito density and hence the baseline transmission intensity in the absence of interventions. It is calculated by

$$K_L = M_0 \frac{2 d_{L_L} \mu_0 \left(1 + d_{P_L} \mu_{P_L}\right) \gamma_L (\lambda + 1)}{\left(\frac{\lambda}{\mu_{L_L} d_{E_L}} - \frac{1}{\mu_{L_L} d_{L_L}} - 1\right)}, \tag{13}$$

where $M_0$ is the initial female mosquito density, $\mu_0$ is the baseline mosquito death rate and

$$\lambda = -\frac{1}{2}\left(\gamma_L \frac{\mu_{L_L}}{\mu_{E_L}} - \frac{d_{E_L}}{d_{L_L}} + \left(\gamma_L - 1\right) \mu_{L_L} d_{E_L}\right) + \sqrt{\frac{1}{4}\left(\gamma_L \frac{\mu_{L_L}}{\mu_{E_L}} - \frac{d_{E_L}}{d_{L_L}} + \left(\gamma_L - 1\right) \mu_{L_L} d_{E_L}\right)^2 + \gamma_L \frac{v \mu_{L_L} d_{E_L}}{2 \mu_{E_L} \mu_0 d_{L_L} \left(1 + d_{P_L} \mu_{P_L}\right)}}, \tag{14}$$

where

$$v = \frac{\beta_L \mu_M e^{-\mu_M / f}}{\mu_M \left(e^{\mu_M / f} - 1\right)\left(1 - e^{-\mu_M / f}\right)}. \tag{15}$$

Here $\beta_L$ is the maximum number of eggs per oviposition per mosquito and

$$\mu_M = -f_R \log\left(p_1 p_2\right), \tag{16}$$

where $p_1$ is the probability of a mosquito surviving one feeding cycle, $p_2$ is the probability of surviving one resting cycle and $f_R$ is the feeding rate.

The model is described by the equations below:

$$\begin{aligned} \frac{dE_L}{dt} &= \beta_L M - \mu_{E_L}\left(1 + \frac{E_L + L_L}{K_L}\right) E_L - \frac{E_L}{d_{E_L}}, \\ \frac{dL_L}{dt} &= \frac{E_L}{d_{E_L}} - \mu_{L_L}\left(1 + \gamma_L \left(\frac{E_L + L_L}{K_L}\right)\right) L_L - \frac{L_L}{d_{L_L}}, \\ \frac{dP_L}{dt} &= \frac{L_L}{d_{L_L}} - \mu_{P_L} P_L - \frac{P_L}{d_{P_L}}. \end{aligned} \tag{17}$$

We assume 50% of the emergent adult mosquitoes are female and all enter the susceptible state $\left(S_M\right)$. These mosquitoes become infected at a rate that depends on the infectiousness of the human population including an appropriate time-lag $\left(\tau_G\right)$ to account for the time taken for parasites to become infectious gametocytes. The force of infection on mosquitoes, $\Lambda_M$, is the sum of the contributions from the different human infection state compartments:

$$\Lambda_M(t)=\frac{\lambda}{\omega}\sum_i\sum_j\zeta_j\psi_i\left(c_D D_{i,j}(t-\tau_G)+c_T T_{i,j}(t-\tau_G)+c_A A_{i,j}(t-\tau_G)+c_U U_{i,j}(t-\tau_G)\right) \quad (18)$$

where $\psi_i$ is the age-dependent biting rate and $\zeta_j$ is the relative biting rate for each heterogeneity compartment defined in equations (2) and (3). The parameter $\lambda$ is the rate at which a person is bitten by mosquitoes, which depends on interventions, and is defined later. The parameter $\omega$ represents a normalising constant for the biting rate over all ages:

$$\omega=\int_0^\infty \psi(a)\mathrm{g(a)da} \quad (19)$$

where $g(a)$ is the human age distribution and is a function of the force of infection for each age group and the proportion of the population in each age compartment at the start of the simulation. The constants $c_U$, $c_D$ and $c_T$ are the infectiousness to mosquitoes from humans in the asymptomatic sub-patent infection, clinical disease and successfully treated compartments and $c_A$ is the infectiousness to mosquitoes from humans in the asymptomatic infection compartment, which is calculated as follows:

$$c_{A_{i,j}}=c_U+\left(c_D-c_U\right)q_{i,j}^{\gamma_1}, \quad (20)$$

where $\gamma_1$ is a fitted parameter for the infectiousness of state *A* and $q_{i,j}$ is the probability of being detected by microscopy, which is given by equation (11).

Once infected, mosquitoes pass through a latent period $\left(E_M\right)$ of fixed length $\tau_M$ and then they become infectious to humans ($I_M$). They are assumed to remain infectious until they die. The infection process in the mosquito population is as follows:

$$\begin{aligned}\frac{dS_M}{dt}&=\frac{P_L}{2d_{P_L}}-\Lambda_M S_M-\mu_M S_M,\\ \frac{\partial E_M}{\partial t}&=\Lambda_M S_M-\Lambda_M(t-\tau_M)S_M(t-\tau_M)P_M-\mu_M E_{M_i},\\ \frac{dI_M}{dt}&=\Lambda_M(t-\tau_M)S_M(t-\tau_M)P_M-\mu_M I_M,\end{aligned} \quad (21)$$

where $\mu_M$ is the mosquito death rate defined in equation (16) and

$$P_M=e^{-\mu_M\tau_M} \quad (22)$$

is the probability that a mosquito survives the extrinsic incubation period. We define the mean EIR experienced by adults at time t to be:

$$\varepsilon_0(t)=\frac{1}{\omega}I_M(t). \quad (23)$$

The parameters for this part of model are given in Table S1.4.

**Table S1.4: Vector model parameters**

| Parameter | Symbol | Estimate |
|---|---|---|
| **Larval model** | | |
| Average number of eggs laid per female mosquito per day | $\beta_L$ | 21.2/day |
| Early instar larval developmental period | $d_{E_L}$ | 6.64 days |
| Late instar developmental period | $d_{L_L}$ | 3.72 days |
| Pupal developmental period | $d_{P_L}$ | 0.643 days |
| Mortality rate of early-stage larvae (density dependent) | $\mu_{E_L}$ | 0.0338/day |
| Mortality rate of late-stage larvae (density dependent) | $\mu_{L_L}$ | 0.0348/day |
| Mortality rate of pupae (density independent) | $\mu_{P_L}$ | 0.249/day |
| Effect of density dependence on late instars relative to early instars | $\gamma_L$ | 13.25 |
| **Infectiousness to mosquitoes** | | |
| Lag from parasites to infectious gametocytes | $\tau_G$ | 12.5 days |
| Untreated disease | $c_D$ | 0.068 |
| Treated disease | $c_T$ | 0.021896 |
| Sub-patent infection | $c_U$ | 0.00062 |
| Parameter for infectiousness of state *A* | $\gamma_1$ | 1.82425 |
| **Mosquito Population Model** | | |
| Baseline daily mortality of adults with no interventions | $\mu_M$ | 0.132 |
| Mean time between feeds | $\delta$ | 3 days |
| Extrinsic incubation period | $\tau_M$ | 10 days |
| Initial female mosquito density | $M_0$ | Dependent on EIR |
| Maximum number of eggs per oviposition per mosquito | $\beta_L$ | 21.2 |
| Probability of surviving one feeding attempt | $p_1$ | Intervention dependent |
| Probability of surviving one resting cycle | $p_2$ | 0.737 |
| Mosquito feeding rate | $f_R$ | Intervention dependent |
| Rate at which a person is bitten by mosquitoes | $\lambda$ | Intervention dependent |

#### Long-lasting insecticide treated nets model

Long-lasting insecticide treated nets (LLIN) have four main effects on the transmission cycle:

i) they increase the overall mosquito death rate;

ii) they lengthen the feeding or gonotrophic cycle;

iii) they change the proportion of bites taken on protected and unprotected people;

iv) they change the proportion of bites taken on humans relative to animals (the Human Blood Index).

We model these impacts on the vector population, as in *Griffin et al.* [1]. The probability of a blood-seeking mosquito successfully feeding depends on the behaviour of the mosquito (which may vary between species) and the anti-vectoral defences employed by the human host population.

As shown in the paper, once a mosquito enters a house to feed, one of three things can happen: it can repeat ($r_N$ ), feed successfully ($s_N$ ) or die ($d_N$). In the full model with both the barrier effect of the LLIN and insecticide, the repellency of LLIN decreases from a maximum, $r_{N0}$, to a non-zero level $r_{NM}$ , at a rate $\gamma_N = \dfrac{\log(2)}{\eta}$ (where $\eta$ is the half-life of the net) reflecting the protection still provided by a net that no longer has any insecticidal effect (and potentially some holes). The killing effect of LLIN decreases from $d_{N0}$ at the same constant rate. Therefore, at a time *t* after nets were distributed,

$$\begin{aligned} r_N &= (r_{N0} - r_{NM})\exp(-t\gamma_N) + r_{NM}, \\ d_N &= d_{N0}\exp(-t\gamma_N), \\ s_N &= 1 - r_N - d_N. \end{aligned} \qquad (24)$$

These values change with the chemicals used as insecticide and the resistance of the mosquitoes to the chemical [5]. We assume a three-yearly distribution of LLINs with adherence to use decaying over time.

The number of mosquitoes entering a house in search of a blood meal can be estimated from experimental hut trials. The presence of a bed net will cause a mosquito to repeat in one of two ways. First, the mosquito will be less likely to enter a house due to the excito-repellent effect of the insecticide on the nets, and secondly once it enters a house it will be repelled from a protected human due to the physical barrier of the net and the effects of the insecticide.

Not all mosquitoes successfully feed upon entering a house even before the introduction of an intervention. Therefore, the probability of repeating, feeding or dying needs to be relative to that seen in the absence of LLINs. The proportion repeating ($r_{N0}$), feeding successfully ( $s_{N0}$ ) and dying ( $d_{N0}$ ) in the presence of LLIN will therefore be,

$$r_{N0} = \left(1-\frac{k_1'}{k_0}\right)\left(\frac{j_1'}{j_1'+l_1'}\right),$$
$$s_{N0} = \frac{k'}{k_0}, \qquad (25)$$
$$d_{N0} = \left(1-\frac{k_1'}{k_0}\right)\left(\frac{l_1'}{j_1'+l_1'}\right),$$

where $j_1' = \left(1-\frac{N_1}{N_0}\right)+\frac{N_1}{N_0}j_1$, $k_1' = \frac{N_1}{N_0}k_1$ and $l_1' = \frac{N_1}{N_0}l_1$. Here, $N_0$ and $N_1$ represent the number of mosquitoes entering a house with or without LLIN. The parameters $j_0$ and $j_1$, $k_0$ and $k_1$, and $l_0$ and $l_1$, denote the percentage of mosquitoes that are not feeding ($j$), are successfully feeding ($k$) or are killed ($l$) in the absence or presence of bed nets. These associations change with resistance. Principally, the mortality effect $r_{N0}$ is reduced and the half-life of nets $\gamma_N$ also reduces [5].

We define the probability of a *Plasmodium falciparum* mosquito biting a human host during a single attempt to be $y$; the probability that a mosquito bites a human host and survives the feeding attempt to be $w$, and the probability of it being repelled without feeding to be $z$. These parameters account for the repeating behaviour observed prior to the introduction of insecticides and exclude natural vector mortality. In this model without indoor residual spraying, $y = w$.

During a single feeding attempt (which may be on animals or humans), a mosquito will successfully feed with probability $W$ depending on the intervention usages given by $c_k$ ($k = 1$ is no intervention compartment and if $k = 2$ is LLIN compartment):

$$W = \sum_{k=1}^{2} w_k c_k, \qquad w_k = \begin{cases} 1 & \text{if } k = 1, \\ 1 - \Phi_b + \Phi_b s_N & \text{if } k = 2, \end{cases} \qquad (26)$$

and be repelled without feeding with probability $Z$ given by

$$Z = \sum_{k=1}^{2} z_k c_k, \qquad z_k = \begin{cases} 0 & \text{if } k = 1, \\ \Phi_b r_N & \text{if } k = 2, \end{cases} \qquad (27)$$

where in both equations $\Phi_b$ is the proportion of bites taken on humans in bed and $s_N$ is the probability of successfully feeding upon an encounter with a net and $r_N$ is the probability of repeating. The average probabilities of mosquitos successfully feeding during a single attempt and being repelled without feeding are:

$$\bar{W} = 1 - Q_0 + Q_0 W, \qquad (28)$$

$$\bar{Z} = Q_0 Z. \qquad (29)$$

The proportion of bites taken on humans in bed is defined as

$$\Phi_B = \frac{\sum_t p_B(t)\lambda_I(t)}{\sum_t \big((1-p_I(t))\lambda_O(t) + p_I(t)\lambda_I(t)\big)},$$

where $\lambda_I(t)$ is the rate at which a person who is indoors at hour *t* is bitten and $\lambda_O(t)$ is the corresponding figure for someone outdoors, and $p_I(t)$ is the proportion of human hosts indoors and $p_B(t)$ in bed at a given time *t.* Due to the lack of data it is assumed that human movement and sleeping patterns are not dependent on age or relative exposure.

The mosquito feeding rate $f_R$ is given by

$$f_R = \frac{1}{\delta_1 + \delta_2}, \tag{30}$$

where $\delta_1$ and $\delta_2$ are the length of time spent looking for a blood meal and resting between feeds respectively. Parameter $\delta_2$ is assumed to be unaffected by the interventions, whilst $\delta_1$ is increased to

$$\delta_1 = \frac{\delta_{10}}{1-\bar{Z}}, \tag{31}$$

where $\delta_{10}$ is the value with no interventions.

The probabilities of surviving the periods of feeding and resting, as mentioned above, are $p_1$ and $p_2$. With no interventions,

$$p_{10} = \exp(-\mu_M \delta_{10}), \tag{32}$$

$$p_2 = \exp(-\mu_M \delta_2), \tag{33}$$

where $\mu_M$ is the baseline mosquito death rate. With interventions $p_2$ is unchanged and

$$p_1 = \frac{\bar{W} p_{10}}{1-\bar{Z} p_{10}}. \tag{34}$$

The probability of surviving one feeding cycle is $p_1 p_2$:

$$p_1 p_2 = \exp(-\mu_M / f_R), \tag{35}$$

hence the mosquito death rate, $\mu_M$, is as defined in equation (16). The probability of surviving the extrinsic incubation period, $P_M$, also changes with intervention usage.

The proportion of successful bites that are on humans depends on the probability of a feeding cycle resulting in a successful bit on a human or an animal. The probability that a feeding cycle ends with a successful bite on a human, $q_H$, is

$$\begin{aligned} q_H &= p_{10}\left(Q_0 W + \bar{Z} q_H\right), \\ q_H &= \frac{p_{10} Q_0 W}{1-\bar{Z} p_{10}}. \end{aligned} \tag{36}$$

The probability that a feeding cycle ends with a successful bite on an animal is,

$$q_A = p_{10}\left(1 - Q_0 + \bar{Z} q_A\right),$$
$$q_A = \frac{p_{10}(1-Q_0)}{1-\bar{Z}p_{10}}. \tag{37}$$

Hence the proportion of successful bites which are on humans is,

$$Q = 1 - \frac{q_A}{q_A + q_H} = 1 - \frac{1-Q_0}{1-Q_0+Q_0 W},$$
$$= 1 - \frac{1-Q_0}{\bar{W}}. \tag{38}$$

and the biting rate on humans is,

$$\alpha = Q f_R. \tag{39}$$

This means that the rate at which a person is bitten by a mosquito is:

$$\lambda_k = \frac{\alpha w_k}{W} \tag{40}$$

and the force of infection of humans on mosquitoes is as defined in equation (18).

Considering different intervention compartments adds a third dimension to the ODEs in equation (1). The EIR now varies according to intervention compartment:

$$\varepsilon_{i,j,k} = \frac{I_M}{\omega} \psi_i \zeta_j \lambda_k. \tag{41}$$

The parameters for this model are given in Table S1.5, how the LLIN parameters vary with pyrethroid resistance are given in **Error! Reference source not found.** and how PBO-LLIN parameters vary with pyrethroid resistance in **Error! Reference source not found.**.

**Table S1.5: LLIN model parameters**

| **Parameter** | **Symbol** | **Estimate** |
|---|---|---|
| Anthropophagy | $Q_0$ | 0.92 |
| Proportion of bites taken on humans in bed | $\Phi_b$ | 0.85 |
| Baseline time spent looking for a blood meal | $\delta_1$ | 0.69 days |
| Time spent resting between feeds | $\delta_2$ | 2.31 days |
